# Neuronal Distribution of Tau Pathology, Microglial Gene Expression Trajectories, and Resilience to Alzheimer disease

**DOI:** 10.64898/2025.12.15.25341752

**Authors:** Sunny Kumar, Ana Claudia Amaral, Cinthya Aguero, Charles Zachary Klein, Michael Siao Tick Chong, Prianca Ramanan, Margaret Elizabeth Scapellato, Riddhimaa Sinha, Julie A Schneider, David A. Bennett, Steven E. Arnold, Matthew P. Frosch, Ibai Diez, Teresa Gomez-Isla

**Affiliations:** Department of Neurology, Mass General Brigham, Boston, MA; Harvard Medical School, Harvard University, Boston, MA; Rush Alzheimer’s Disease Center, Rush University Medical Center, Chicago, IL; C.S. Kubik Laboratory for Neuropathology, Mass General Brigham, Boston, MA, USA; Center for Inflammation Imaging, Mass General Brigham, Boston, MA; Computational Neuroimaging Lab, Biobizkaia Health Research Institute, Barakaldo, Spain; IKERBASQUE Basque Foundation for Science, Bilbao, Spain

**Keywords:** Alzheimer disease, tau pathology, neurites, synapses, microglia, gene expression, resilience

## Abstract

**Importance:** Some individuals are capable of tolerating Alzheimer disease neuropathological changes (ADNC) without manifesting clinical symptoms. Elucidating the neuropathological, and molecular mediators may facilitate the identification of more accurate *in vivo* biomarkers and inform the development of targeted therapeutic strategies.

**Objective:** To investigate cellular distribution of tau pathology, microglial responses, and gene expression profiles associated with divergent clinical outcomes (dementia vs. no dementia) in individuals exhibiting comparable ADNC.

**Design, Setting and Participants:** We analyzed postmortem brain tissue from 97 participants from the ROSMAP study:49 with high likelihood of AD (25 demented [demented AD] and 24 cognitively normal [resilient]), and 48 with low likelihood (22 demented [impaired-other (IMP-O)] and 26 cognitively normal [control]). Cases were matched for age, gender, and co-pathologies.

**Main outcomes and measures:** Amyloid-β plaques, phospho-tau pathology (tangles and tau-positive neurites), tau oligomers in synaptic-fractions, tau seeding activity, neurons, synapse density, and astrocyte and microglia activation were quantified. The relationships with bulk and microglia-specific RNA-sequencing were also examined. Statistical analyses employed ANOVA with Tukey’s HSD/Bonferroni corrections for pathological and clinical variables, and the Wald test for differential gene expression

**Results:** Demented AD and resilient brains exhibited comparable tau tangle and amyloid-β plaques; however, demented AD showed higher total pTau burden (tangles and tau-positive neurites) (Mean[SD], 27.39%[21.89%] vs. 10.60%[14.71%]; p= 0.0004) and elevated pTau oligomer levels in synaptic-enriched fractions (0.48[0.52] vs. 0.16[0.19]; p=0.0010). Both demented AD (0.21×10^7^[0.14×10^7^]; p<0.0001) and IMP-O (0.62×10^7^ [0.42×10^7^]; p<0.0001) showed greater synaptic loss than resilient (1.54×10^7^[0.55×10^7^]; p=0.0008) and controls (2.92×10^7^[1.13×10^7^]). CD68+ microglia burden was increased in demented (1.14%[0.27%]; p<0.0001) and IMP-O (0.97%[0.36%]; p<0.0001) but not in resilient (0.72% [0.17%]; p=0.2835) compared to controls (0.59%[0.17%]). Synaptic pTau oligomers and CD68+ microglia were the strongest correlates with antemortem cognition. Resilient brains exhibited downregulation of neuroinflammation-related genes and possessed a distinct microglial subpopulation supporting resilience, characterized by overexpression of CD83, DUSP1, and NAMPT.

**Conclusion and relevance:** Our findings suggest that aberrant accumulation of tau in neurites and synapses, rather than tangles within neuronal soma, may trigger a microglial pro-inflammatory activation linked to synaptic loss and impaired cognition. Cell-specific transcriptomic analysis identified a distinct microglial cell subpopulation associated with resilience to ADNC.

## Introduction

About one-third of elderly individuals harbor in their brain classic Alzheimer disease neuropathologic changes (ADNC) (e.g., plaques and tangles) at autopsy, which would be expected to have had devastating clinical consequences but never developed dementia [1–4]. We and others have termed this phenomenon ‘resilience’ to ADNC [2, 3, 5–8]. Understanding the underlying mechanisms is likely to provide key insights for developing novel biomarkers and therapeutic strategies to prevent cognitive decline in the setting of plaques and tangles. Our previous studies demonstrated better preservation of neurons, synaptic markers, and axonal geometry in resilient compared to demented AD [2, 5, 9]. Integrity of dendritic spine density [10] and cortical thickness [11] has also been consistently associated with resilience to ADNC [12, 13]. We recently applied expansion microscopy (ExM), a novel technique that enables 4-5 fold physical magnification of brain tissue [14–17], to investigate the spatial relationships among glial cells, tau oligomers, and individual synaptic elements in the human brain. Our data suggested that the abnormal presence of tau oligomers in synapses may serve as the ‘eat-me’ signal, triggering excessive engulfment of synapses by microglia and astrocytes in demented AD brains, leading to early loss of brain function [9]. A minimal accumulation of tau oligomers in synapses of resilient brains was associated with suppressed glial inflammatory responses and better preservation of synapses and cognition [9]. Those studies provided important initial insights into potential mechanisms underlying human brain resilience to ADNC, but the absence of longitudinal antemortem cognitive assessments in some of the cases was a key limitation. Here, we had the unique opportunity to conduct detailed neuropathological and biochemical assessments combined with bulk and single-nucleus RNA-sequencing data analyses in an informative cohort of 97 brains from participants in the Religious Orders Study and the Rush Memory and Aging Project (ROSMAP) [18] representing four categories (‘demented AD’, ‘resilient’, ‘impaired-other (IMP-O)’, and ‘control’; demented AD and resilient were at equivalent stages of tau tangle and amyloid plaque pathology at autopsy) to gain further insight into the neuropathological, cellular, and molecular mediators of brain resilience to ADNC. All ROSMAP participants agree to annual detailed cognitive assessments and brain donation after death. This study addresses three main questions: 1) Is the presence of tau pathology within specific neuronal compartments (e.g. soma, neurites, synapses) associated with distinct microglial responses and divergent fate of synapses and cognition between demented AD and resilient, despite comparable burdens of ADNC at autopsy? 2) Do IMP-O brains without overt ADNC, Lewy body, TDP-43, vascular, or other defined pathologies to meet criteria for any primary neuropathologic diagnosis also exhibit a pro-inflammatory microglial activation linked to synaptic loss and impaired cognition? 3) Are there specific microglial subpopulations promoting brain resilience to ADNC?

## Material and methods

### Human Brain Samples

This postmortem study included 97 brains from ROSMAP participants. All ROSMAP participants enrolled without known dementia and agreed to annual clinical evaluation and brain donation. Both studies were approved by an Institutional Review Board of Rush University Medical Center and all participants signed informed and repository consents and an Anatomic Gift Act. The mean interval between the last cognitive assessment and death was 11 [18] (months). Autopsies were performed according to standardized protocols [19]. Demographic, cognitive, and neuropathological characteristics of the study sample are summarized in **Table 1**. Brains were divided in 4 groups according to National Institute on Aging and Reagan Institute (NIA-RI) neuropathologic criteria [20]: the first group comprised brains from 25 cognitively impaired individuals fulfilling criteria for high likelihood of AD [‘demented AD’]; the second group comprised brains from 24 cognitively normal individuals fulfilling criteria for high likelihood of AD [‘resilient’]; the third group comprised brains from 22 cognitively impaired individuals fulfilling criteria for low likelihood of AD [‘IMP-O’] and no overt ADNC, Lewy body, TDP-43, vascular, or other defined lesions sufficient for any primary neuropathologic diagnosis; and the fourth group comprised brains from 26 cognitively normal individuals fulfilling criteria for low likelihood of AD [‘controls’]. The four groups were matched for age, sex, years of education, other brain co-morbidities (cerebrovascular lesions, Lewy bodies, and TDP-43 aggregates), and post-mortem intervals (PMIs). Demented AD and resilient were also matched for Braak neurofibrillary tangle stage and CERAD neuritic plaque score. This study followed the Strengthening the Reporting of Observational Studies in Epidemiology (STROBE) reporting guidelines.

**Table 1.** Summary of the demographic characteristics of the postmortem brain samples. Continuous values are represented as mean (standard deviation). * p-value <0.05 compared to controls; ** p-value <0.005 compared to controls; ^+^ p-value <0.05 compared to demented AD; ^++^p-value <0.005 compared to demented AD; PMI, post mortem interval (hours); CERAD neuritic plaque score: A neuropathologic diagnosis based on semiquantitative measure of neuritic plaques (CERAD; C0: none – C1:Sparse – C2: Moderate – C3: Frequent) [3]; NIA-Reagan, presence of AD relying on both neurofibrillary tangles (Braak) and neuritic plaques (CERAD) [83, 84]; CAA, semiquantitative summary of cerebral amyloid angiopathy (CAA) pathology (0:none; 1 mild; 2 moderate; 3 severe) in 4 neocortical regions: midfrontal, midtemporal, parietal, and calcarine cortices [85]; Atherosclerosis rating was made by visual inspection (0:none; 1 mild; 2 moderate; 3 severe)[86]; Lewy body disease, describes 4 stages of distribution of a-synuclein in the brain (0-No present; 1-niagral predominant; 2-limbic type; 3-neocortical type)[87]; TDP-43, presence of TDP-43 cytoplasmic inclusions in neurons and glia(None TDP-43 pathology or restricted to the amygdala – TDP-43 pathology extending beyond the amygdala)[88]; MMSE, Mini-Mental State Examination. Nineteen different cognitive tests were used to create the 3 different memory composites [89]: Episodic memory included 7 memory tests (immediate and delayed recall of the East Boston Story and Story A from Logical Memory, Word List Memory, Word List Recall, Word List Recognition); language/semantic memory included 3 tests (15-item Boston Naming Test, Verbal Fluency, 15-item word reading test); and working memory included 3 tests (Digit Span Forward, Digit Span Backward, Digit Ordering). Visuospatial ability was computed as a composite of 2 test: line orientation, and progressive matrices (16 items). Psychomotor speed was computed as a composite of 2 test: line orientation, and progressive matrices (16 items). Composite measures provided by RUSH study were constructed by converting raw scores on component tests to z scores, using the baseline mean and SD of all persons in the parent studies, and then averaging the component z scores to yield the composite score. All the previous tests were used to compute the global cognitive score.

|  | Control<br>[n=26] | Impaired, other<br>[n=22] | Resilient<br>[n=24] | Demented AD<br>[n=25] |
| --- | --- | --- | --- | --- |
| **Demographics** |  |  |  |  |
| Age (years) | 90.55 (5.42) | 87.63 (6.50) | 91.03 (5.19) | 90.58 (5.52) |
| Sex (% Females) | 58% | 50% | 54% | 56% |
| Years of educ | 17.08 (3.69) | 16.32 (3.22) | 16.58 (3.22) | 16.24 (3.22) |
| APOE4+ | 3 (12%) | 4 (18%) | 6 (25%) | 11 (44%) * |
| **Neuropathology** |  |  |  |  |
| Brain weight | 1,218 (117) | 1,161 (112) | 1,153 (110) * | 1,128 (133) * |
| PMI (h) | 7.22 (3.14) | 7.53 (5.77) | 8.37 (4.52) | 6.97 (4.41) |
| Braak NFT stage | 0(1), I(8), II(10),<br>III(7) | I(5), II(7), III(10)** | V (24) ** | V (24), VI (2) ** |
| CERAD score | 13 – 9 – 4 – 0 | 13 – 5 – 4 – 0** | 0 – 0 – 10 – 14** | 0 – 0 – 7 – 18** |
| NIA-Reagan | Low (26) | Low (22)** | Mid(10),<br>High(14)** | Mid(6), High(19)** |
| Diffuse plaques | 0.56 (0.94) | 0.12 (0.30)** | 1.22 (0.87)* | 1.32 (0.80)** |
| CAA | 0.96 (0.79) | 0.86 (0.94)** | 1.46 (0.83)* | 1.8 (0.76) ** |
| Microinfarcts | 2 | 7* | 6 | 4 |
| Atherosclerosis | 0 (8), 1(13), 2(3),<br>3(2) | 0(4), 1(10), 2(7), 3(1) | 0 (6), 1(12), 2(6) | 0(5), 1(12), 2(5),<br>3(3) |
| Lewy body disease | None | None | 2 limbic | None |
| TDP-43 | 20 – 6 | 11 – 10 | 13 – 10 | 7 – 15** |
| **Cognition** |  |  |  |  |
| MMSE | 28.07 (1.57) | 16.81 (8.65) **** | 27.62 (1.74) ** | 11.02 (9.53) ** |
| Episodic Memory | 0.50 (0.41) | -1.60 (1.03) **** | 0.22 (0.47) ** | -2.49 (0.98) ** |
| Visuospatial Ability | 0.15 (0.69) | -1.22 (1.04) ** | 0.08 (0.62) ** | -1.32 (1.13) ** |
| Psychomotor Speed | -0.29 (0.59) | -1.97 (0.96) ** | -0.55 (0.84) ** | -2.18 (0.98) ** |
| Language/Semantic | 0.16 (0.43) | -1.36 (1.38) ** | 0.05 (0.27) ** | -1.92 (1.55) ** |
| Working Memory | 0.25 (0.70) | -1.46 (0.74) ** | -0.32 (0.40) **** | -1.58 (1.14) ** |
| Global Cognition | 0.26 (0.29) | -1.61 (0.73) ** | -0.04 (0.32) **** | -2.19 (1.03) ** |

### Neuropathological and Biochemical Assessments

We conducted quantitative neuropathological assessments of amyloid-β (Aβ) plaques, tau pathology (neurofibrillary tangles (NFTs) and tau positive neurites), neurons, and activated astrocytes and microglial cells in the superior temporal sulcus (STS) and dorsolateral prefrontal cortex (dlPFC). Expansion microscopy (ExM), a technique enabling physical expansion of tissue by 4– to 5-fold, was used to quantify synaptic elements. Western Blot was used to measure pTau (monomers and oligomers) in synaptosome-enriched fractions, and in vitro tau seeding activity assays were conducted. See Supplementary Methods for protocols, antibody details (eTable 1), and quantitative approaches.

### Transcriptional and Single-Cell Analysis

We analyzed available bulk and single-nucleus RNA-sequencing (snRNA-seq) data from the dlPFC of a subset of the ROSMAP participants included in this study. RNA-seq was used for Differential Gene Expression and subsequent Gene-Set Enrichment Analyses to identify biological processes associated with resilience (14 demented AD, 14 resilient, 17 IMP-O, 21 control). snRNA-seq data was utilized to determine microglial subpopulation proportions and perform trajectory analysis to model gene expression changes along microglial state trajectories (8 demented AD, 9 resilient, 5 IMP-O, 11 controls). Data retrieval, analytical pipelines, and workflows are detailed in the Supplementary Methods.

### Statistical Analyses

Statistical analyses were performed using GraphPad Prism v9.4.1. Data were tested for normality and homogeneity of variance. One-way ANOVA or its non-parametric equivalent was used for group comparisons, followed by Tukey’s or Bonferroni’s post hoc tests. Correlation analyses were conducted using the Spearman test. A two-sided Wilcoxon signed-rank test was used to compare cell proportions between groups.

## Results

### **β**-Amyloid Plaque Deposition and Tau Aggregation in Neurofibrillary Tangles and Neurites

We quantified burdens of total and compact β-amyloid plaques in the STS, defined as percentage of cortex occupied by Aβ plaques labeled by 6F3D antibody and Thioflavin-S staining, respectively (eFigure 1a), number of NFTs labeled by PHF-1 antibody, and total phospho-Tau (pTau) burden, defined as percentage of cortex occupied by PHF-1 positive NFTs and neurites (Figure 1a). Neither total plaque burden (mean [SD]; 3.40% [1.80%] in demented AD vs. 2.49% [2.06%] in resilient; p=0.7843) nor Thioflavin-S positive plaque burden (0.82% [0.75%] vs. 0.60% [0.31%]; p>0.9999) differed significantly between demented AD and resilient (eFigure 1a). Number of NFTs showed no differences either between demented AD and resilience (259.40 [189.40] vs 139.40 [154.40]; p>0.9999). Likewise, total plaque burden (5.03% [3.15%] in demented AD vs. 3.49% [2.49%] in resilient; p>0.9999) and NFT counts (48.26 [65.48] vs. 15.83 [31.26]; p=0.8325) in dlPFC did not differ between demented AD and resilient (not shown). However, demented AD showed a significantly higher total pTau burden (PHF-1 positive NFTs plus neurites) compared to resilient (27.39% [21.89%] vs. 10.60% [14.71%]; p=0.0004) (Figure 1a).

**Figure 1:**
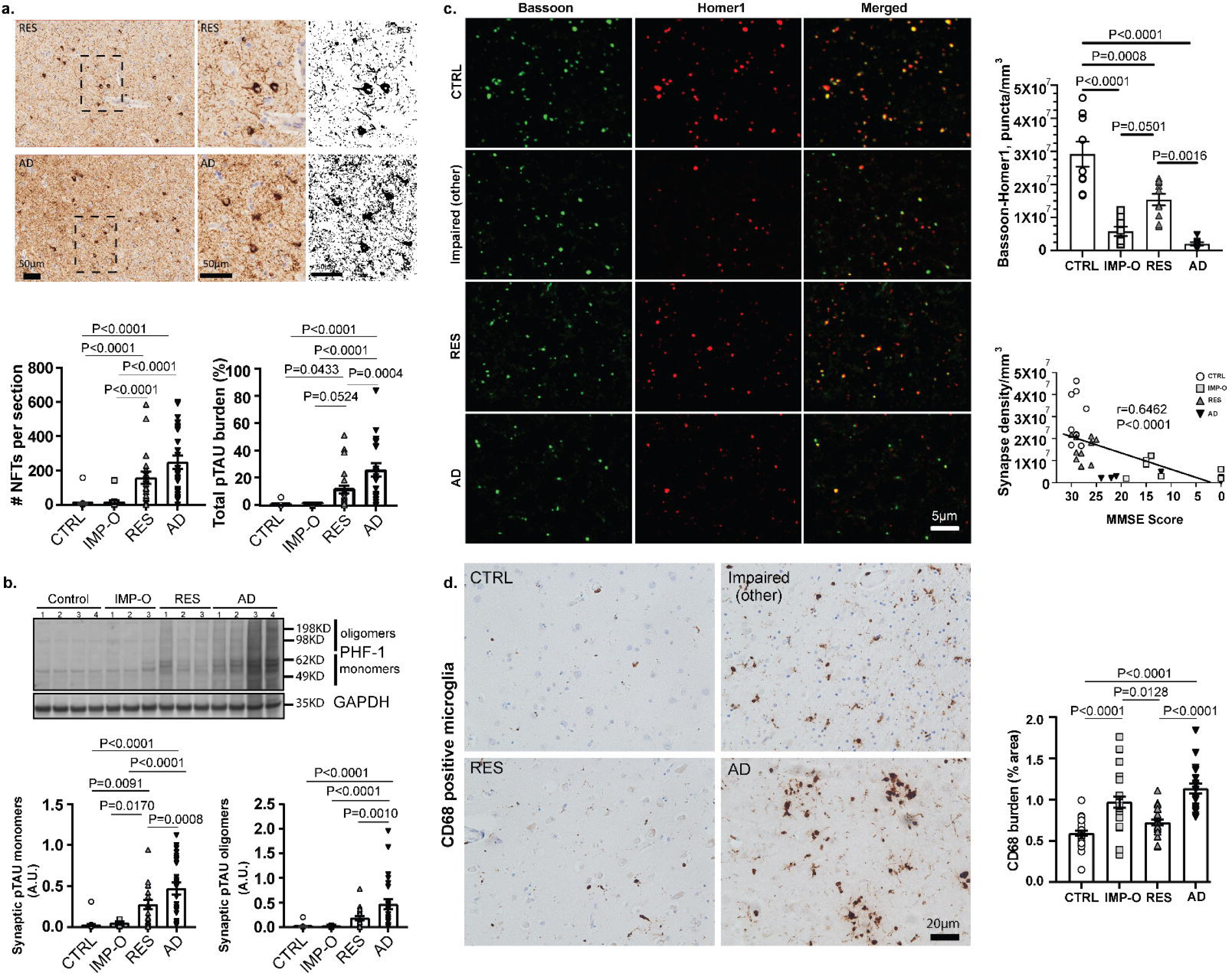
(**a**) Representative images and quantification of PHF1 immunostaining in the STS of control (0-II), IMP-O (0-II), Resilient (Braak V-VI), and Demented AD (Braak V-VI). Demented AD showed comparable numbers of NFTs but higher total pTau-burden (tangles plus pTau positive neurites) compared to resilient. **(b)** Representative images and quantification by Western Blot of pTau-PHF1 levels (monomeric and oligomeric) in STS synaptosome enriched fractions. Resilient showed significantly lower levels of pTau monomers and oligomers compared to demented AD. **(c)** Representative images of pre-synapse (Bassoon; Green), post-synapse (Homer; Red), and colocalizing (Bassoon and Homer) synapse densities in the STS; quantification of colocalizing (Bassoon and Homer) synapse densities; and correlation of synaptic density with MMSE scores. Synapse densities were significantly reduced in demented AD and IMP-O compared to resilient and controls. **(d)** Representative images and quantification of CD68+ microglia burden in the STS. CD68+ burden was significantly increased in demented AD and IMP-O but not in resilient compared to controls.

These findings suggest that pTau pathology burden in neurites may impact cognitive outcomes more than classic NFTs localized in the neuronal soma.

### Tau Hyperphosphorylation and Oligomerization in Synaptosome-Enriched Fractions

We assessed levels of tau hyperphosphorylation (PHF-1 antibody) and oligomerization by WB analyses of synaptosome-enriched fractions. Levels of monomeric and oligomeric pTau were significantly higher in synaptosome-enriched fractions of demented AD compared to resilient (mean [SD]; monomer 0.48 [0.36] vs. 0.23 [0.24]; p=0.0008; oligomers 0.48 [0.52] vs. 0.16 [0.19]; p=0.0010) (Figure 1b). Negligible levels of pTau monomers and oligomers were present in IMP-O and controls (monomer 0.03 [0.02] and 0.03 [0.06]; oligomer 0.01 [0.01] and 0.01 [0.04]) (Figure 1b).

These findings suggest that aberrant accumulation of hyperphosphorylated monomeric and oligomeric tau in synapses may be a key pathological alteration linked to divergent synaptic fates and cognitive outcomes between demented AD and resilient.

### Tau Seeding Activity

Tau seeding activity in the STS did not significantly differ between demented AD and resilient (mean [SD]; 99.02 [75.07] vs. 56.97 [44.95]; p>0.9999) (eFigure 2a,2b). Of note, tau seeding activity in demented AD and resilient strongly correlated with NFT number (R=0.6262, p<0.0001) and total pTau burden (R=0.5355, p<0.0001) (eFigure 2c,2d).

**Figure 2:**
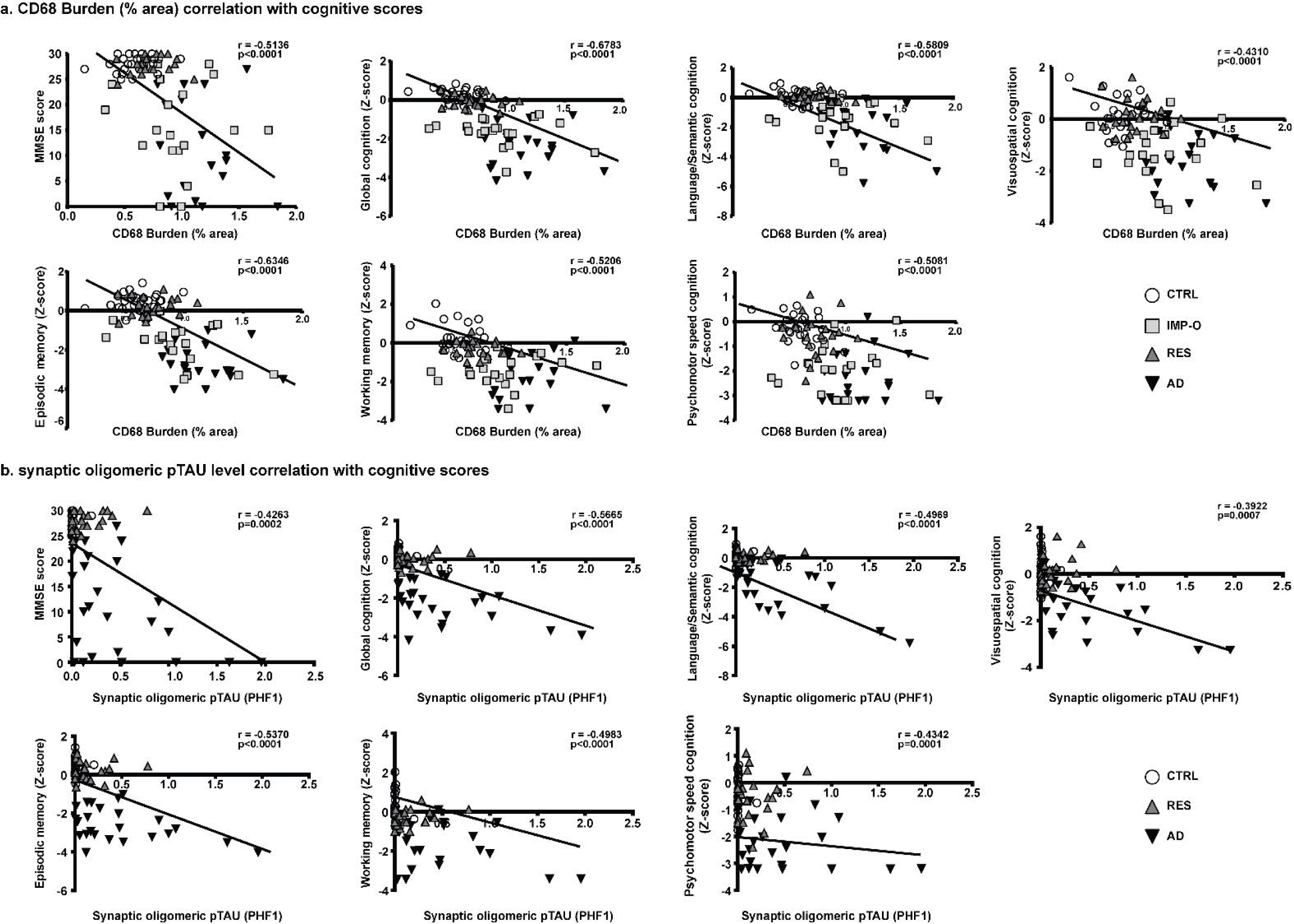
Representative graphs depicting the correlation analyses between pathological and ante-mortem cognitive measures in control (Braak 0-II), IMP-O (0-II), Resilient (Braak V-VI), and Demented AD (Braak V-VI) **(a)** CD68+ burden and **(b)** synaptic oligomeric pTau levels in the STS significantly inversely correlated with MMSE scores, global cognitive performance, and specific cognitive domains, including language/semantic, visuospatial, episodic memory, working memory, and psychomotor speed.

These results suggest that tau seeding activity is primarily driven by burden of pTau aggregates, and unlikely to explain the different cognitive outcomes between these two groups.

### Neuronal counts, Synaptic densities, and Astrocyte and Microglial Responses

We quantified number of neurons and density of colocalizing pre– and post-synaptic (mature) puncta in the STS of demented AD, resilient, IMP-O, and control brains. Number of neurons was significantly lower in demented AD (mean [SD]; 303.7 [73.45]; p=0.0212) but not in resilient (342.10 [63.56]; p>0.9999) or IMP-O (350.40 [107.40]; p>0.9999) compared to controls (374.7[56.46]) (eFigure 1c).

Density of mature puncta was significantly lower in demented AD (0.21×10^7^ [0.14×10^7^]; p<0.0001), resilient (1.54×10^7^ [0.55×10^7^]; p=0.0008), and IMP-O (0.62×10^7^ [0.42×10^7^]; p<0.0001) compared to controls (2.92×10^7^ [1.13×10^7^]), but resilient exhibited a significantly higher number of mature puncta compared to demented AD (p=0.0016) and IMP-O (p=0.0501) (Figure 1c). Significant negative correlations were observed between mature synapse densities and ante-mortem Mini-Mental State Examination (MMSE) scores (R=-0.6462; p<0.0001) (Figure 1c). Analyses of demented AD and resilient revealed significant correlations between mature synapse density and total pTau burden (PHF-1 positive NFTs plus neurites); (R=-0.5130; p=0.0074), and between mature synapse density and levels of oligomeric pTau (R=-0.4061; p=0.0355) in synaptosome-enriched fractions (eFigure 3a,3b).

**Figure 3:**
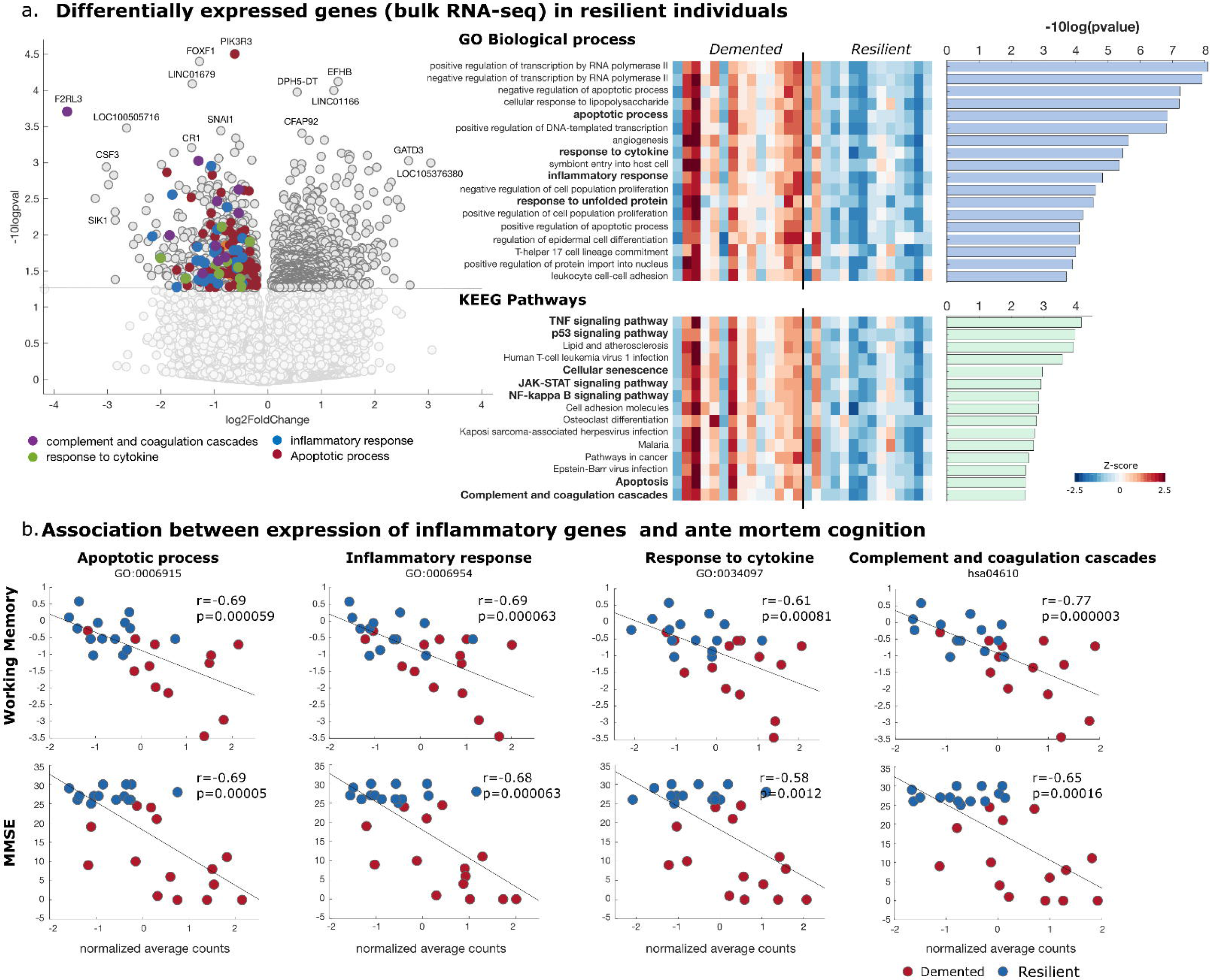
Bulk RNA sequencing data. **(a)** Volcano plot displaying differentially expressed genes in resilient individuals compared to demented AD. Gene set-enrichment of differentially expressed genes (p-value <0.05) for GO biological processes and KEEG pathways. A normalized aggregate expression matrix clearly shows the reduced expression of the different annotations. **(b)** The correlation of biological processes (apoptotic process, inflammatory response, response to cytokine, and complement and coagulation cascade) revealed a negative association with antemortem cognitive measures (MMSE score and working memory).

We also measured burdens of GFAP+ astrocytes and CD68+ microglial cells in STS. We found a significantly higher burden of activated astrocytes (GFAP+) in demented AD (31.58% [16.14%]; p=0.0006) but not in resilient (20.04% [7.07 %]; p=0.9007) or IMP-O (20.45 % [10.38%]; p=0.8646) compared to controls (17.71% [8.70 %]) (eFigure 1d). CD68+ microglial burden was significantly higher in demented AD (1.14% [0.27%]; p<0.0001) and IMP-O (0.97% [0.36%]; p<0.0001) but not in resilient (0.72% [0.17%]; p=0.2835) compared to controls (0.59% [0.17%]) (Figure 1d). Mature synapse density negatively correlated with both CD68+ microglial burden (R=-0.3575, p=0.0524; eFigure 3c) and GFAP+ astrocyte burden (R=-0.3330, p=0.0625) but narrowly failed to reach statistical significance.

These findings suggest that pro-inflammatory activation of microglial cells may impair synaptic integrity, whereas reduced activation in resilient may contribute to the relative better preservation of synapses despite similarly high plaque and tangle burdens. The elevated microglial pro-inflammatory response in IMP-O brains, even without overt β-amyloid or tau pathology, further supports a deleterious role of microglial activation on synaptic integrity and brain function.

### Clinico-Pathological Correlations in Demented AD and Resilient

CD68+ microglial burden and pTau oligomers in synapses emerged as two of the strongest correlates with multiple antemortem cognitive measurements. We found significant negative correlations between CD68+ microglial burden and antemortem cognitive scores for MMSE (R=-0.5136; p<0.0001), global cognition (R=-0.6783; p<0.0001), language/semantic (R=-0.5809; p<0.0001), visuospatial (R=-0.4310; p<0.0001), episodic memory (R=-0.6346; p<0.0001), working memory (R=-0.5206; p<0.0001), and psychomotor speed (R=-0.5081; p<0.0001) (Figure 2a).

Measures of oligomeric pTau in synaptosome-enriched fractions also showed significant negative correlations with MMSE (R=-0.4263; p=0.0002), global cognition (R=-0.5665; p<0.0001), language/semantic (R=-0.4969; p<0.0001), visuospatial (R=-0.3922; p=0.0007; Figure 2b), episodic memory (R=-0.5370; p<0.0001), and working memory (R=-0.4983; p<0.0001) and psychomotor speed (R=-0.4342; p=0.0001) (Figure 2b). Total pTau burden (PHF-1 positive NFTs plus neurites) was significantly correlated with CD68+ microglial burden (R=0.4639; p≤0.0001).

These findings suggest that aberrant accumulation of pTau pathology in neurites and synapses, rather than classic tau tangles in the neuronal soma, is linked to the pro-inflammatory response of microglia, potentially underlying the divergent fates of synapses and cognition observed between demented AD and resilient.

### Transcriptomic Signatures of Brain Resilience to ADNC

We performed gene-set enrichment analysis of differentially expressed genes in demented AD compared to resilient. Using bulk RNAseq data from dlPFC, we identified 631 downregulated genes in resilient compared to demented brains (p-value<0.05; Figure 3a). GO biological process enrichment revealed significant downregulation of pathways related to apoptotic process, inflammatory response, and response to cytokines, among others, in resilient compared to demented AD (Figure 3a, eFigure 4,5). Enrichment of KEGG pathways revealed downregulation in TNF, p53, and JAK-STAT and NF-kappa B signaling pathways, and the complement and coagulation cascades. No significant upregulated GO biological processes and KEEG pathways were found in resilient compared to demented AD. Expression changes of significant genes for each annotation are shown in eFigure 4, 5. While global AD pathology burden (a ROSMAP provided aggregate measure derived from plaques and tangles across five brain regions) showed modest correlation with antemortem working memory (R=0.44; p=0.0232), aggregated gene expression values of significant biological pathways showed stronger associations to apoptotic process (R=0.69; p<0.0001), inflammatory response (R=0.60; p<0.0001), response to cytokines (R=0.61; p=0.0008), and complement and coagulation cascades (R=0.77; p<0.0001) (Figure 3b, eTable 2). Associations with other cognitive domains are shown in eTable2.

**Figure 4:**
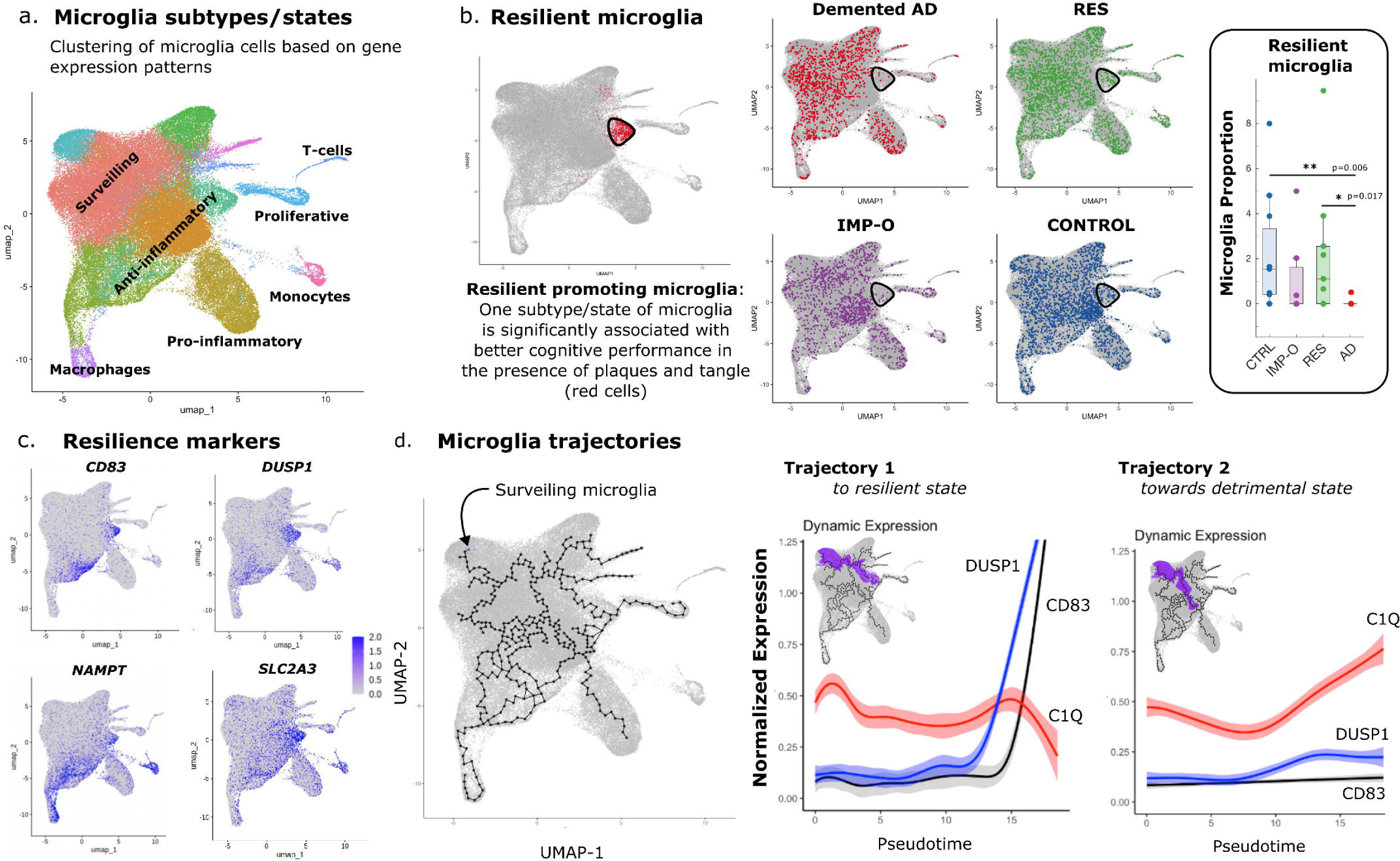
Microglia State Promoting Resilience. **(a)** Single-nucleus RNA sequencing data was used to cluster cells based on their transcriptomic signatures. Known marker genes were used to assign each cell to one of seven major cell types (excitatory and inhibitory neurons, astrocytes, oligodendrocytes, microglia, oligodendrocyte-promoting cells, and vascular niche). **(b)** Microglial cells were further subclustered in different states based on their transcriptomic profiles. The proportion of a specific microglial subpopulation was significantly associated with resilience and control individuals (plotted in red). Cells from each group were displayed in different colors (demented: red, resilient: green, IMP-O: purple, controls: blue). **(c)** The expression of four markers characteristic of this microglial subtype is shown. **(d)** Microglia trajectory analysis identified diverse pathways through which microglia can transition between functional states. The trajectory characterized by the progression from a surveillant state to a resilience-promoting state was marked by a distinct pattern: an increase in resilience markers accompanied by a downregulation of complement C1q. This pattern contrasted with the trajectory observed when microglia transitioned to a more detrimental state.

Additionally, IMP-O brains showed upregulation of genes associated with neuronal action potential, glutamatergic and dopaminergic synaptic transmission, and nicotine and morphine addiction compared to controls (eFigure 7). Conversely, we identified 280 genes commonly downregulated in both IMP-O and AD versus controls, enriched for metabolism related processes (including metabolic pathways and fatty acid metabolism/elongation) and erythrocytes O2/CO2 exchange. Compared to controls, IMP-O also showed downregulation of genes in the TNF signaling pathway.

### Resilience Promoting Microglial Cell Subpopulation

Cell proportion analysis revealed a significantly higher abundance of a specific microglial cell subpopulation in resilient and control relative to demented AD (p=0.0177 and p=0.0059; Figure 4c). A trend toward a decrease in this same microglial cell subpopulation was observed in IMP-O, although the difference did not reach statistical significance (p=0.1402). This ‘resilient’ microglial cell subtype was characterized by overexpression of genes such as CD83, DUSP1, NAMPT, and SLC2A3 among others, with key roles in resolving inflammation and preventing excessive inflammatory responses (eTable 3 for a complete list of overexpressed genes). Trajectory analysis comparing the progression of microglial cells from a surveillance state to ‘resilient’ state vs. a detrimental state revealed distinct patterns: as expression of ‘resilient’ microglial cell markers (CD83, DUSP1, NAMPT, and SLC2A3) increased, expression of the complement C1q marker decreased, suggesting a shift toward a more beneficial microglial cell state (Figure 4d) that may explain the better preservation of synapses and neurons in resilient compared to demented AD.

## Discussion

This cross-sectional study assessed a cohort of human brains from individuals with extensive antemortem longitudinal cognitive assessments as part of their participation in the ROSMAP [18] before autopsy. Brains were classified into four categories: 1) demented with high probability of AD (demented AD), 2) non-demented with high probability of AD (resilient), 3) demented with low probability of AD (IMP-O), and 4) non-demented with low probability of AD (controls). Demented AD and resilient were matched for tau Braak stages and CERAD neuritic plaque scores but had widely divergent antemortem cognitive statuses (dementia vs. preserved cognition). The aims of this study were twofold: 1) to investigate neuronal compartment-specific patterns (soma, neurites, synapses) of tau pathology and their relationship to microglial responses and synaptic loss, and 2) to gain mechanistic insights into differential gene expression signatures linked to divergent microglia responses that might account for the different outcomes of synapses and cognition between demented AD and resilient.

Previous studies showed that robust burdens of ADNC (e.g., Aβ plaques and neurofibrillary tangles) can be present at autopsy in some individuals without antemortem cognitive impairment, underscoring that these two lesions are not sufficient to cause dementia in all cases, highlighting the phenomenon of human brain resilience [2, 3, 5–8]. Numerous studies, including our own, demonstrated that number of NFTs is more strongly associated with disease duration and clinical progression in AD [21–23] compared to amyloid plaque burden [1, 22, 23]. However, neuronal loss in AD brains far exceeds the number of NFTs observed at autopsy [22], a finding supported by studies in mice demonstrating that tangle-bearing neurons are long-lived, dying at lower rates than neighboring non-tangle-bearing neurons [24, 25]. These paradoxical observations suggest that other forms of pathological tau, distinct subcellular localizations, and/or non-tau related mechanisms may play a more direct role in driving neurodegeneration in AD. The contributions of soluble versus aggregated tau species [26–28], as well as their compartment-specific localization within neurons (e.g., soma, neurites, synapses) [9, 29–31], to neuronal and synaptic loss and cognitive impairment remain incompletely understood. Prior studies by our group and others demonstrated aberrant accumulation of soluble high molecular weight hyperphosphorylated tau (e.g. oligomers) within synapses of demented AD brains [2, 31, 32], with markedly reduced levels in resilient brains [2, 9]. The present study expands on these findings in an independent cohort and further reveals a strong correlation between aberrant accumulation of pTau in neurites and synapses with synaptic loss and antemortem cognitive measures. Importantly, pTau accumulated at substantially lower levels in neurites and synapses of resilient despite comparably high NFT burdens, suggesting that neuritic and synaptic tau lesions likely exert a more pronounced detrimental effect on neurons and synapses than classic NFTs.

Synapse loss is a hallmark feature of AD and one of the earliest and most robust pathological correlates of cognition [5, 9, 33]. Understanding synaptic vulnerability is critical for developing preservation strategies to halt cognitive decline in AD [34–36]. Our previous studies showed that oligomeric tau accumulation at synapses seemed to act as an “eat-me” signal linked to an excessive glial-mediated synaptic engulfment in demented AD [9]. This mechanism was attenuated in resilient brains and associated with better preservation of synapses, suggesting a potential link between reduced oligomeric tau-driven synaptic clearance and brain resilience to ADNC. In the present study, we also identified a significant reduction in synaptic density in IMP-O brains in the absence of ADNC and no primary neuropathologic diagnosis, implicating AD-independent processes in their synaptic vulnerability and cognitive impairment. These observations prompted us to conduct a more in-depth examination of the phenotypic profiles of glial cells across the different brain groups.

In response to accumulating ADNC, both microglia and astrocytes undergo marked phenotypic changes characterized by a reactive activation state [37–39]. While glial activation may initially serve protective functions, chronic and dysregulated responses are increasingly implicated in exacerbating neurodegeneration and synaptic integrity loss [9, 33, 40]. Aligned with previous findings [2, 41], in the current study we observed significant increases of activated astrocytes (GFAP+) and microglia (CD68+) in demented AD but not in resilient. This attenuated pro-inflammatory glial response in resilient likely supports better preservation of their cognitive function and synaptic integrity. Intriguingly, in IMP-O brains, while the burden of activated astrocytes remained comparable to controls, activated microglial burden was significantly elevated, supporting the notion that microglial activation may also contribute to synaptic loss and cognitive impairment even in the absence of overt ADNC. Importantly, other work has also demonstrated that microglial contributes to cognitive decline, independent of major neuropathologies [42].

The strongest correlates of cognitive performance in this cohort were neither amyloid plaques nor neurofibrillary tangles, but rather pTau burden in synapses (in cases with ADNC) and activated microglial burden (in all cases with or without overt ADNC).

Building on our detailed quantitative pathological assessments, we pursued deeper mechanistic insights by integrating available transcriptomic data. Such analyses enable identification of molecular pathways and cell subpopulations linked to ADNC vulnerability versus resilience [43]. We observed robust downregulation of genes associated with inflammatory processes in resilient compared to demented AD, which was strongly correlated with antemortem cognitive performance in a dose-dependent manner. Consistent with our pathological data, these findings suggest that reduced complement activity may favorably influence clinical outcomes, potentially via downregulation of microglia-mediated synapse engulfment.

Single-cell transcriptomic studies have identified distinct gene expression signatures in subsets of neurons [44–47] and astrocytes [46, 48] associated with better cognitive performance and resilience to ADNC. In the present study, we focused on microglia, as they emerged as a common denominator linked to synaptic loss. We identified a distinct microglial subtype that promotes resilience against ADNC characterized by the expression of genes such as CD83, DUSP1, NAMPT, and SLC2A3, among others. Of note, a previous study identified depletion of this microglial cluster in AD, multiple sclerosis, and severe COVID-19, suggesting that this microglial state may represent a shared neuroprotective mechanism across diverse neuroinflammatory and neurodegenerative pathologies [49].

CD83 expression has been associated with early microglial activation in AD as well as rapid resolution of neuroinflammation [50, 51]. Its deficiency/deletion in murine models results in over-activated microglia and impaired inflammatory resolution [50, 51]. Similarly, DUSP1 may confer neuroprotection in AD by suppressing amyloid-β generation, deactivating stress-activated MAPKs, and reducing neuroinflammation and neuronal apoptosis [52–55]. NAMPT is a key metabolic and immunomodulatory enzyme declining with aging and AD, and its loss impairs mitochondrial function and worsens inflammation [56, 57]. NAMPT confers neuroprotection in stroke and neurodegeneration by limiting neuroinflammation (promoting anti-inflammatory glia, reducing leukocyte infiltration, and preserving BBB integrity) [58] and by supporting neuronal survival via NAD⁺/SIRT1/AMPK signaling and astrocyte-derived exosomal NAMPT–triggered AMPK/mTOR autophagy [59, 60]. Post-mortem studies also show consistent reductions in SLC2A3 (GLUT3) in AD brains (reviewed in [61]). Interestingly, recent preclinical work in tauopathy models indicates that increasing GLUT3 expression can suppress inflammatory responses and reduce tau-associated degeneration [62]. In this context, a resilient promoting microglia population might increase the expression of genes such as CD83, DUSP1, and NAMPT to limit and control the inflammatory response, while genes such as GLUT3 might reflect a metabolic adaptation to maintain the microglia’s increased energetic demands imposed by ADNC.

The resilient-promoting microglial state identified here was also significantly enriched for genes associated with interleukins 4 and 13 signaling. These interleukins limit neuroinflammation by promoting anti-inflammatory, pro-regenerative microglial states that protect neurons [63, 64].

Our previous study also observed increased levels of IL-4 and IL-13 in the entorhinal cortex of resilient brains [65]. IL-4 upregulation exerts neuroprotective effects by promoting anti-inflammatory phenotypes and limiting apoptosis [66–73]. Similarly, peripheral IL-13 administration [74, 75] enhances plasticity, memory, and learning [76], and neuronal survival in mouse CNS injury models [77]. In vitro and in vivo murine studies also show that IL-4 and IL-13 enhance amyloid-β clearance and improve cognitive performance [78–81]. Together, these data suggest that the microglia state identified in the present study may promote resilience to ADNC by restraining pro-inflammatory responses and mitigating microglia-mediated synaptic damage. Of note, this resilient microglial subtype was decreased in both demented AD and IMP-O brains, further favoring its potentially broader role in safeguarding synapses and thereby preserving cognition in human brains beyond the context of ADNC.

Intriguingly, the transcriptomic analysis of IMP-O brains showed no upregulation of inflammatory genes but revealed hyperexcitability in glutamatergic and dopaminergic neurons, along with decreased metabolism and TNF signaling. Thus, microglial dysfunction may underlie hyperexcitability in IMP-O brains, as microglia normally restrain neuronal activity [82]. Increased CD68^⁺^ microglial burden in IMP-O brains occurred without a corresponding rise in GFAP^⁺^ reactive astrocytes, suggesting that chronic microglial dysfunction alone may disrupt synapses and thus cognition in the absence of ADNC. Additional factors such as chronic stress, infections, addiction, and metabolic disturbances may contribute, and further studies are needed to define the cellular and molecular mechanisms underlying this IMP-O phenotype.

In conclusion, tau pathology in neurites and synapses strongly correlated with pro-inflammatory activation of microglia and marked loss of synapses in demented AD brains. In contrast, resilient brains showed reduced pathological tau in neurites and synapses despite similar burdens of ADNC, with suppressed microglial activation and better preservation of synapses. Notably, elevated microglial cell pro-inflammatory activation also occurred in IMP-O brains without ADNC and was associated with synaptic loss and impaired cognition. Trajectory-based gene expression analyses further identified, for the first time in human brains, a distinct microglial cell subpopulation linked to resilience to ADNC.

### Study Limitations

Postmortem human brain studies offer critical insights but are inherently cross-sectional, capturing only a single time point. As such, they reveal associations but cannot establish causality or temporal dynamics of disease processes and thus require cautious interpretation.

## Supporting information

Manuscript

## Data Availability

All data produced in the present study are available upon reasonable request to the authors

## Acknowledgments

The results published here are in part based on data obtained from the AD Knowledge Portal (https://adknowledgeportal.org). Study data were provided by the Rush Alzheimer’s Disease Center, Rush University Medical Center, Chicago. We are deeply indebted to all the volunteers in the Rush Religious Order Study, and the staff engaged in subject assessment, autopsy, and brain banking at Rush University Medical Center. Data collection was supported through funding by NIA grants P30AG10161 (Bennett) and P30AG72975 (Schneider), (ROS), R01AG15819 (ROSMAP; genomics and RNAseq), R01AG17917 (Bennett) (MAP and RNAseq), R01AG36836 (RNAseq), and the Illinois Department of Public Health (Bennett) (ROSMAP). Additional phenotypic data can be requested at www.radc.rush.edu.” snRNAseq data collection was funded by NIH grants U01AG061356 (De Jager/Bennett), RF1AG057473 (De Jager/Bennett), and U01AG046152 (De Jager/Bennett) as part of the AMP-AD consortium, as well as NIH grants R01AG066831 (Menon) and U01AG072572 (De Jager/St George-Hyslop). S.K. was supported by Alzheimer’s Association Research Fellowship (AARF-23-1150672).

S.E.A. was supported by NIH R01AG039478 grant. I.D. was supported by Alzheimer’s Association (AARF-23-1145358), Fundación ADEY, and the Spanish Ministry of Science (RYC2022-035429-I and PID2023-150633OA-I00). T.G.I. was supported by P30AG062421 and Cure Alzheimer’s Fund grant.

## Supplementary Material

**eMethods**.

**eReferences**.

**eTable 1.** Antibodies Utilized in the Present Study

**eTable 2**. Correlation of cognitive measures with GO biological function and KEGG pathway

**eTable 3.** List of upregulated genes in resilient promoting microglia

**eFigure 1**. Quantification of β-amyloid plaque burden, neuronal density, and GFAP-positive astrocyte burden in the STS

**eFigure 2**. Tau seeding assays and analyses examining relationships with NFT counts and total pTau burden in the STS

**eFigure 3**. Correlation analyses of synapse density with total pTau burden, synaptic pTau oligomer levels, and CD68+ microglia burdens in the STS

**eFigure 4**. Genes implicated in the regulation of altered biological processes in demented AD compared to resilient brains

**eFigure 5**. Genes implicated in dysregulated pathways in demented AD compared to resilient brains

**eFigure 6**. Workflow for cell type annotation in snRNAseq and identification of microglia cells

**eFigure 7**. Differential gene expression of IMP-O compared to control brains

