## Supplementary material for "Neuronal Distribution of Tau Pathology, Microglial Gene Expression Trajectories, and Resilience to Alzheimer disease": Manuscript

**eMethods.**

**eReferences.**

**eTable 1.** Antibodies Utilized in the Present Study

**eTable 2**. Correlation of cognitive measures with GO biological function and KEGG pathway

**eTable 3.** List of upregulated genes in resilient promoting microglia

**eFigure 1.** Quantification of β-amyloid plaque burden, neuronal density, and GFAP-positive astrocyte burden in the STS

**eFigure 2.** Tau seeding assays and analyses examining relationships with NFT counts and total pTau burden in the STS

**eFigure 3.** Correlation analyses of synapse density with total pTau burden, synaptic pTau oligomer levels, and CD68+ microglia burdens in the STS

**eFigure 4.** Genes implicated in the regulation of altered biological processes in demented AD compared to resilient brains

**eFigure 5.** Genes implicated in dysregulated pathways in demented AD compared to resilient brains

**eFigure 6.** Workflow for cell type annotation in snRNAseq and identification of microglia cells

**eFigure 7.** Differential gene expression of IMP-O compared to control brains

### eMethods

#### 1a. Immunohistochemical Staining and Quantitative Pathological Analysis

We conducted quantitative assessments of amyloid-β (Aβ) plaques, tau inclusions (neurofibrillary tangles (NFTs) and tau positive neurites), neurons, synapses, and activated astrocytes and microglial cells following our previously published protocols [1-3] and appropriate antibodies. The antibodies utilized in the present study are summarized in the supplementary eTable 1. Analyses were performed on 6 μm-thick paraffin-embedded brain sections containing the superior temporal sulcus (STS), a multisensory cortical region involved in integration of auditory, visual, and tactile information, and the dorsolateral prefrontal cortex (dlPFC), involved in higher-order executive functions. Both regions are known to be consistently affected by Aβ plaque and neurofibrillary tangle deposition in AD [1].

Post-primary antibody incubation, sections were treated with appropriate horseradish peroxidase-conjugated secondary anti-mouse or anti-rabbit antibodies (BOND Polymer Refine Detection, catalog number DS9800, Leica Biosystems Newcastle, UK). Post-secondary antibody incubation, immunolabels were visualized using 3,3'-diaminobenzidine (DAB). Sections were counterstained with hematoxylin as per the Leica Biosystems auto-stainer protocol. Post-staining, slides were dehydrated and mounted in a Paramount mounting medium. Slides were scanned at 20X objective for capturing images with a Nanozoomer XR (NDP.scan 3.3.3, Hamamatsu Photonics, Hertfordshire, UK).

Total amyloid-β plaque burden, defined as the percentage of the superior temporal sulcus (STS) per section occupied by 6F3D-immunostained plaque deposits, compact amyloid-β plaque burden, defined as the percentage of the superior temporal sulcus per section occupied by thioflavin-S positive amyloid-β plaques, and the number of neurofibrillary tangles (NFTs) immunostained with PHF-1 antibody per section were quantified following our published protocols [2, 4]. Total pTAU burden (tangles plus pTau positive neurites) in the STS per section was quantified using the QuPATH analysis platform. Burdens of GFAP-positive astrocytes and CD68-positive microglia in the STS per section were quantified following our previously published protocol [2]. HucD-positive neurons in the STS per section were quantified using the semi-automatic deep-learning-based artificial intelligence AIFORIA platform following our previously published method [5]. Total amyloid-β plaque burden and number of NFTs were also quantified in the dlPFC as describe above.

#### 1b: Expansion Microscopy (ExM)

Expansion microscopy (ExM) enables isotropic physical expansion of tissue specimens in three dimensions by a factor of approximately 4- to 5-fold, allowing optical resolution of individual synaptic elements using conventional microscopy [3, 6, 7]. ExM was performed according to our previously published protocols [3] with minor modifications (eMethods 1b in Supplemental Figure). We achieved a mean (SD) tissue expansion factor of 4.49 (0.48), consistent with values previously reported by us and others [3, 7-9] .

The ExM procedure involves multiple steps for the preparation of human paraffin-embedded brain tissue sections for synaptic analysis as described in our previously published protocol [5] and is briefly summarized below.

1. **Deparaffinization:** Formalin-fixed, paraffin-embedded brain sections (6μm thick) from the STS region were pre-heated at 68°C for 30 minutes. The sections were deparaffinized using xylene (twice for 5 minutes) and rehydrated through an ethanol gradient (from 100%-95%-90%-80%-50%-ddH2O (2 x 5mins).

2. **Anchoring:** A water-soluble AcX stock solution (5mg/500µl in DMSO) was diluted (1:100 in PBS) and applied to the tissue sections for 3-4 hours at room temperature in a humidified environment. The sections were then washed (PBS 2X5 minutes) and dried.
3. **Gelling:** A fresh gelling solution was freshly prepared and applied to the sections (For 500 µl gelling solution, 467.5 µl of monomer solution (2.5% acrylamide, 8.6% sodium acrylate, and 0.1% N-N-methylene bis-acrylamide, diluted in distilled water), 10 µl of 4HT, 10 µl of TEMED, and 12.5 µl of APS were added in this sequential order. A gel chamber was created using a coverslip (24x40mm, Thermo Fisher, catalog number 102440), and the gel was incubated overnight in a humidified environment.
4. **Digestion:** The gel is trimmed around the tissue after removing the coverslip. The trimmed gels, still mounted on the glass slide, were placed in 50ml Falcon tubes. Digestion buffer was prepared by adding beta-mercaptoethanol (0.7%) to the digestion solution (5% 1M Tris, 5% 500mM EDTA, 0.5% Triton-X, 20% SDS, diluted in distilled water). The digestion step was performed in an autoclave at 121°C for 60 mins. The samples were left to cool down inside the autoclave for at least 3-6 hours.
5. **ExM Gel Processing:** The gels were removed, washed with PBS (3X5 minutes), and were then cut into small sections (~0.5x0.5 cm) and stored in a 24-well plate (Greiner Bio-One, Fisher Scientific, catalog number 07000680) for further analysis.
6. **Immunostaining of ExM Gels**: Immunostaining was conducted within glass bottom 24-well plates. ExM gels were blocked briefly using MaxBlock solution (Active Motif) for 2 hours at room temperature, followed by 3 x 1-minute washes in MaxWash buffer (Active Motif). Primary antibodies were prepared at a 1:100 dilution in MaxStain solution (Active Motif) and incubated with the ExM gels for 72 hours at 4°C. Post-incubation, ExM gels were washed 4x10 minutes each in MaxWash buffer before incubating overnight at 4°C with secondary antibodies (Alexa 488, Alexa 555) diluted 1:100 in MaxStain solution. The gels were subsequently washed five times for 10 minutes each in MaxWash buffer and then incubated 3x10 minutes in distilled water (dd$H_2O$). The expansion factor of the ExM gels was measured, and for long-term storage, the gels were placed in PBS and stored at 4°C.

Expanded tissue sections were imaged using an Olympus FV3000 confocal microscope (resolution, 1024 × 1024 pixels). A z-stack spanning 0.40 µm was acquired to optimize discrimination of true signal (defined as synaptic puncta present in at least 2 consecutive optical sections) from potential artifacts [10]. For synapse density measures, 4 to 6 fields of view per case were randomly selected in cortical layers II-III of the STS. Immunostaining for Bassoon and Homer1 was used to label presynaptic and postsynaptic elements, respectively. Quantification of colocalized presynaptic and postsynaptic 'mature' puncta was performed using Imaris software (Bit-Plane). Individual layers for presynaptic and postsynaptic elements were created separately by masking the background and setting up a diameter of the signal sphere of 0.4 µm. Presynaptic and postsynaptic layers were independently masked, and colocalization analysis was restricted to puncta within a surface distance threshold of 0.276 µm, excluding voxels beyond this range.

**1c: Protein Extraction, Synaptosome Preparation, and Immunoblotting**

Frozen tissue containing the STS was used for total, cytosolic, and synaptosome fraction preparation following our previously published protocol [2, 4] . In brief, tissue was homogenized using a Teflon-glass mechanical tissue grinder in Buffer A solution (25mM HEPES 7.5, 120 mM NaCl, 5mM KCL, 1mM MgCl2, 2mM CaCl2, 1mM DTT) containing Phosphatase Inhibitor Cocktail Tablets (Roche, 04906845001), and Protease Inhibitor Cocktail tablets (Roche,

11697498001). After homogenization, the extract was filtered through two 80 µm Millipore Nylon filters to purify the total extract. For this, 200 µL of homogenate was mixed with 200 µL of distilled water and 70 µL of 10% SDS, then passed three times through a 27½ G needle. The homogenate was then boiled for 5 minutes, centrifuged it at 15,000 g for 15 min, and the total extract supernatant was collected. For extracting synaptosomes, we passed the remaining homogenates through PALL Acro disc Syringe 5µm Filters and centrifuged at 1000g for 10 minutes at 4°C. Post-centrifugation, the supernatant was separated and ultracentrifuged at 100,000g for 45 minutes to pellet synaptosomes. The supernatant was collected and used as the cytosolic fraction. The pellet was then resuspended in 200µL Buffer B (50mM Tris, 1.5% SDS, 1mM DTT), boiled for 5 minutes, and centrifuged at 15,000 g for 15 min. The supernatant was collected and used as the synaptosome fraction.

Once fractions were purified, we measured the protein concentration using a BCA reagent (Sigma, USA). 20–30 µg of protein samples were loaded and separated using MES SDS Running Buffer (Novex, NP0002) and 4–12% Bis-Tris Novex gels (Invitrogen, MAN0003679). Post-transfer, the nitrocellulose membranes were blocked for an hour at room temperature using Odyssey Blocking Buffer (LiCor, 927-40000). Membranes were probed with PHF1 (antibody kindly donated by Dr. Peter Davies) and human tau (Dako, A0024) primary antibodies. GAPDH (Millipore, AB2302) was used as the housekeeping protein for normalization. Licor secondary antibodies (IR Dye 680RD donkey anti-chicken 926-68075, IR Dye 800 CW Donkey anti-rabbit 926-32213, IR Dye 800CW Donkey anti-mouse 926-32212) were then used to visualize bands using the Odyssey Infrared Imaging System (V3.0). ImageStudio was used to quantify the bands of interest by drawing rectangles of an equal area around each band of interest. The background was corrected by taking the median of the area three pixels to the left and right of each region of interest and subtracting that value from the measured signal.

**1d: Tau Seeding Activity Assays**

We conducted tau seeding activity assays using PBS fractions from fresh frozen brain homogenates containing the STS isolated from individual cases following our previously published protocols [11, 12]. The tau seeding activity assays were conducted in vitro using a stably cyan fluorescent protein (CFP) TauRDP301S and yellow fluorescent protein (YFP) TauRDP301S (Tau RD P301S FRET Biosensor (ATCC CRLR-3275™)) expressing HEK293 cell line [11, 12]. In brief, HEK293 cells were plated at 40,000 cells/well in black clear-bottom 96-well plates coated with poly-D-lysine (Corning). The following day, the cells were treated with a total of 50 µl solution containing 2 µg/well PBS-soluble frozen brain lysate (3,000 g) and 7% lipofectamine 2000 transfection reagent (11668–019, Invitrogen) in Opti-MEM (11058–021, Life technologies). Before the treatment, lysates and lipofectamine complexes were incubated at room temperature for 20 min. For the treatment protocol, cells were incubated for 41 h with the lysate-lipofectamine complex at 37°C. Post-treatment, cells were collected using Trypsin-EDTA and moved into another 96-well U-bottom plate (Corning) containing chilled DMEM 10% FBS media. Using a plate spinner, cells were pelleted twice at 1,800 g and fixed with 2% paraformaldehyde (PFA, 15714-S, Electron Microscopy Sciences) in the dark for 10 min at 4 °C. Cells were then resuspended immediately in cold PBS and run on the MACSQuant VYB flow cytometer (Miltenyi). The HEK293 cells were excited with a 405 nm laser, and then the CFP and FRET signals were read using 405/50 nm and 525/50 nm filters, respectively. We used MACS Quantify flow cytometry software (Miltenyi) to calculate the integrated FRET density (IFD) by multiplying the % number of FRET-positive tau aggregates by the mean fluorescence intensity of FRET-positive tau aggregates. The resulting values were normalized to the basal FRET signal obtained from Lipofectamine-treated cells (without brain lysate), which served as the untreated, FRET-negative control population. All samples were run in triplicates, and representative images of the tau aggregates were captured using the ZOE Fluorescent Cell Imager (BioRad).

**1e: Differential Gene Expression Analysis**

We retrieved RNA-seq data from dorsolateral prefrontal cortex (dlPFC) of ROSMAP participants from the AMP-AD Knowledge Portal in accordance with all terms and conditions (Synapse IDs: syn22283382; syn2230160) [13, 14]. The dataset included raw transcript counts for 60,603 genes from 14 demented AD, 14 resilient, 17 IMP-O, and 21 control brains used for neuropathological and biochemical analyses in the present study. Differential gene expression analysis was performed using the DESeq2 R package [15]. Transcript's read counts were modeled using a negative binomial distribution and a generalized linear model to fit the experimental condition. The model included the four groups (demented AD, resilient, IMP-O, and controls) and relevant confounding factors: age, sex, sequencing batch, and PMI. A Wald test was used to identify differentially expressed genes between demented AD and resilient. To visualize gene counts and perform correlation with cognitive variables, we extracted normalized counts of each gene using the variance-stabilizing transformation available in DESeq2.

To identify biological processes and pathways associated with resilience to ADNC, gene-set enrichment analyses were performed using the NIH Database for Annotation, Visualization, and Integrated Discovery (DAVID) platform [16]. All differentially expressed genes with a p-value < 0.05 were included. Enrichment results were assessed with respect to Gene Ontology (GO) biological processes [16] and Kyoto Encyclopedia of Genes and Genomes (KEGG) pathways. Only annotations that remained significant following multiple testing correction (FDR q < 0.05) are reported. For each significant GO process and KEGG pathway, we calculated an aggregate score by averaging normalized expression levels of genes significantly differentially expressed between demented AD and resilient within each annotation. Aggregate scores were then used to correlate with cognitive variables and to visualize expression patterns.

**1f: Preprocessing of snRNA-seq data**

To gain initial insight into the microglia molecular mechanism promoting resilience to ADNC, we used available snRNA-seq data on nuclei isolated from frozen dlPFC tissue samples of 489 ROSMAP participants (Synapse IDs: syn16780177; syn51123521). Only participants with RIN > 5 and PMI < 24 hours were included. We used Seurat V5 workflow [17] to process snRNAseq data and identify microglial cells (eMethods 1e in Supplemental Figure 6). dlPFC snRNA-seq data were available on 33 (including 8 demented AD, 9 resilient, 5 IMP-O, and 11 controls) of the 97 brains used for neuropathological and biochemical studies and were used to generate a reference single nucleus RNAseq dataset [18]. We used Seurat V5 workflow [17] to normalize and scale the data for each cell. Then, we identify the 3,000 genes with the highest cell-to-cell variation in the dataset and perform dimensionality reduction using principal component analysis (PCA) to retain 30 components. We then applied the Louvain community detection algorithm to a k-nearest neighbors graph of nuclei to detect different groups of cells. The 172,659 available cells were divided into 16 distinct clusters, and we matched each cluster to one of seven major cell types using previously reported cell type markers [18-20]: excitatory neurons, inhibitory neurons, astrocytes, microglia (including monocytes and macrophages), oligodendrocytes, OPCs, and vascular niche cells (including vascular cells, pericytes, SMCs, and fibroblasts). The cells of the other 420 individuals [21] were mapped to this reference dataset using reference-based reciprocal PCA integration, and each cell was assigned to one of the major 7 cell types. UMAP embedding was used to visualize the clustering and evaluate the harmonization procedure.

**1g: Microglia sub-states**

For each library, we performed sub-clustering of microglia cells using the same steps as for whole snRNA-seq preprocessing. Then, for each cluster, we removed low-quality cells - those cells that were outliers (three interquartile distances above the third quartile) for the total number of UMIs, number of unique genes or mitochondrial RNA. We also used DoubletFinder [22] to detect and eliminate doublets; clusters with 70% or more doublets were also removed. Then, we performed the integration/alignment between libraries using Seurat anchor-based CCA integration. Once the data was integrated, we performed clustering to identify different microglia subgroups based on their expression using different resolutions (Lovain algorithm over a k-nearest neighbors' graph of nuclei). The n=87,739 cells classified as microglia were used to delineate state diversity and subpopulations. Established markers were used to assign functional annotations to each state/cluster [18, 23-25]. For each brain, we calculated the proportion of each microglia subpopulation relative to total number of microglia. Of the 97 brains studied neuropathologically and biochemically, snRNA-seq data were available for 33 (8 demented AD, 9 resilient, 5 IMP-O, and 11 controls). A two-sided Wilcoxon signed-rank test compared microglia subtype proportions between groups. Marker genes for each microglia subpopulation were identified via Seurat differential expression analysis against remaining microglial cells.

We used Monocle 3 [26] to perform trajectory analysis of microglial cells, enabling the reconstructing sequential gene expression changes and identifying branching points for cell fate decisions. Two trajectories from a surveillance state were examined: one leading to a resilient state and another to a pro-inflammatory state. A pseudotime value was assigned to each cell and key marker dynamics modeled along those trajectories.

**eTable1.** Antibodies Utilized in the Present Study

| Antibody | Host, clonality | IHC/IF | ExM | WB | Company | Cat. Number |
|---|---|---|---|---|---|---|
| Aß (6F3D) | Mouse, monoclonal | 1:600 | | | Dako | M0872 |
| pTAU (PHF1) | Mouse, monoclonal | 1:500 | 1:100 | 1:1000 | Davies | N/A |
| Bassoon | Rabbit, polyclonal | | 1:100 | | Synaptic Systems | 141 003 |
| Homer1 | Guinea pig, polyclonal | | 1:100 | | Synaptic Systems | 160 004 |
| GAPDH | Rabbit, monoclonal | | | 1:1000 | Cell Signaling | 14C10 |
| GFAP | Mouse, monoclonal | 1:20000 | | | Sigma Aldrich | G3893 |
| CD68 | Mouse, monoclonal | 1:500 | | | Dako | M0814 |
| Alexa Fluor 488 | Goat | | 1:100 | | Invitrogen | A10436 |
| HuCD | Mouse, monoclonal | 1:200 | | | Millipore | MABN153 |
| Alexa Fluor 555 | Goat | | 1:100 | | Invitrogen | A30677 |

**eTable 2**. Correlation of cognitive measures with GO biological function and KEGG pathway.
** p-value <0.05; ** p-value <0.005; *** p-value <0.0005;*

| Terms | Episodic Memory | Visuospatial ability | Perceptual Speed | Semantic Memory | Working Memory | Global Cognition | MMSE |
|---|---|---|---|---|---|---|---|
| **GO Biological Process** | | | | | | | |
| positive regulation of transcription by RNA polymerase II | -0.62** | -0.53** | -0.43* | -0.52* | -0.69*** | -0.68*** | -0.72*** |
| negative regulation of transcription by RNA polymerase II | -0.63*** | -0.47* | -0.48* | -0.49* | -0.69*** | -0.66*** | -0.72*** |
| negative regulation of apoptotic process | -0.51* | -0.42* | -0.33 | -0.32 | -0.67*** | -0.55** | -0.63*** |
| cellular response to lipopolysaccharide | -0.56** | -0.44* | -0.42* | -0.47* | -0.66*** | -0.62** | -0.64*** |
| apoptotic process | -0.59** | -0.46* | -0.42* | -0.43* | -0.69*** | -0.62** | -0.69*** |
| positive regulation of DNA-templated transcription | -0.60** | -0.48* | -0.40* | -0.43* | -0.72*** | -0.64*** | -0.68*** |
| angiogenesis | -0.49* | -0.37 | -0.37 | -0.39* | -0.66*** | -0.52* | -0.63*** |
| response to cytokine | -0.48* | -0.36 | -0.33 | -0.37 | -0.61** | -0.53** | -0.58** |
| symbiont entry into host cell | -0.50* | -0.33 | -0.37 | -0.34 | -0.64*** | -0.51* | -0.62*** |
| inflammatory response | -0.55** | -0.50* | -0.39* | -0.50* | -0.69*** | -0.63*** | -0.68*** |
| negative regulation of cell population proliferation | -0.58** | -0.52* | -0.42* | -0.48* | -0.77*** | -0.65*** | -0.69*** |
| response to unfolded protein | -0.35 | -0.35 | -0.22 | -0.15 | -0.58** | -0.39* | -0.56** |
| positive regulation of cell population proliferation | -0.52* | -0.38* | -0.3 | -0.36 | -0.66*** | -0.55** | -0.63*** |
| positive regulation of apoptotic process | -0.61** | -0.51* | -0.38 | -0.44* | -0.70*** | -0.65*** | -0.71*** |
| regulation of epidermal cell differentiation | -0.48* | -0.48* | -0.40* | -0.42* | -0.69*** | -0.57** | -0.58** |
| T-helper 17 cell lineage commitment | -0.46* | -0.28 | -0.36 | -0.38 | -0.48* | -0.51* | -0.54** |
| positive regulation of protein import into nucleus | -0.49* | -0.3 | -0.35 | -0.26 | -0.65*** | -0.48* | -0.58** |
| leukocyte cell-cell adhesion | -0.35 | -0.18 | -0.24 | -0.25 | -0.51* | -0.38* | -0.46* |
| **KEEG Pathways** | | | | | | | |
| TNF signaling pathway | -0.56** | -0.39* | -0.38 | -0.45* | -0.63*** | -0.60** | -0.65*** |
| p53 signaling pathway | -0.65*** | -0.44* | -0.40* | -0.42* | -0.59** | -0.64*** | -0.68*** |
| Lipid and atherosclerosis | -0.48* | -0.33 | -0.39* | -0.35 | -0.64*** | -0.53** | -0.61*** |
| Human T-cell leukemia virus 1 infection | -0.56** | -0.35 | -0.46* | -0.40* | -0.57** | -0.59** | -0.64*** |
| Cellular senescence | -0.60** | -0.37 | -0.44* | -0.46* | -0.57** | -0.62** | -0.65*** |
| JAK-STAT signaling pathway | -0.56** | -0.38* | -0.41* | -0.44* | -0.59** | -0.60** | -0.64*** |
| NF-kappa B signaling pathway | -0.52* | -0.34 | -0.37 | -0.39* | -0.57** | -0.55** | -0.58** |
| Cell adhesion molecules | -0.53** | -0.31 | -0.39* | -0.34 | -0.58** | -0.54** | -0.56** |
| Osteoclast differentiation | -0.58** | -0.33 | -0.46* | -0.48* | -0.46* | -0.59** | -0.65*** |
| Kaposi sarcoma-associated herpesvirus infection | -0.49* | -0.32 | -0.40* | -0.42* | -0.62** | -0.55** | -0.59** |
| Malaria | -0.36 | -0.17 | -0.27 | -0.25 | -0.51* | -0.40* | -0.48* |
| Pathways in cancer | -0.60** | -0.48* | -0.40* | -0.49* | -0.70*** | -0.66*** | -0.67*** |
| Epstein-Barr virus infection | -0.50* | -0.29 | -0.40* | -0.32 | -0.51* | -0.52* | -0.59** |
| Apoptosis | -0.51* | -0.32 | -0.33 | -0.31 | -0.62** | -0.53** | -0.62*** |
| Complement and coagulation cascades | -0.58** | -0.45* | -0.44* | -0.47* | -0.77*** | -0.61** | -0.65*** |
| Growth hormone synthesis, secretion and action | -0.62** | -0.44* | -0.46* | -0.47* | -0.72*** | -0.65*** | -0.66*** |
| C-type lectin receptor signaling pathway | -0.52* | -0.41* | -0.44* | -0.50* | -0.64*** | -0.61** | -0.64*** |

**eTable 3.** List of upregulated genes in resilient promoting microglia. Genes whose expression is predominantly restricted to resilience-promoting microglia subtype in bold.

| **Gene** | **pval** | **avg_log2FC** | **pct.1** | **pct.2** | **pval_adj** | | **Gene** | **pval** | **avg_log2FC** | **pct.1** | **pct.2** | **pval_adj** |
|---|---|---|---|---|---|---|---|---|---|---|---|---|
| **RGS1** | 0 | 2.31541072 | 0.724 | 0.263 | 0 | | CDKN1A | 2.97E-102 | 1.83443322 | 0.265 | 0.106 | 1.09E-97 |
| **SRGN** | 0 | 2.59470702 | 0.81 | 0.373 | 0 | | AL118516.1 | 3.89E-101 | 2.37536928 | 0.158 | 0.046 | 1.42E-96 |
| **FOS** | 0 | 2.86941978 | 0.556 | 0.129 | 0 | | AL512603.2 | 2.40E-96 | 3.38849646 | 0.061 | 0.009 | 8.80E-92 |
| **SLC2A3** | 0 | 2.92752957 | 0.632 | 0.22 | 0 | | HIST1H2BF | 5.04E-96 | 2.47427013 | 0.111 | 0.027 | 1.84E-91 |
| **DUSP1** | 0 | 2.47876501 | 0.537 | 0.142 | 0 | | MYOCOS | 1.58E-94 | 3.47649355 | 0.055 | 0.008 | 5.78E-90 |
| AC131944.1 | 0 | 2.45113414 | 0.642 | 0.259 | 0 | | HIST1H3J | 2.53E-93 | 3.52117002 | 0.043 | 0.005 | 9.27E-89 |
| HIST1H2AC | 0 | 2.47644658 | 0.645 | 0.305 | 0 | | GADD45B | 1.37E-87 | 2.36225502 | 0.136 | 0.039 | 5.02E-83 |
| **DDIT4** | 0 | 2.51239708 | 0.485 | 0.149 | 0 | | GLUL | 9.56E-87 | 1.1721526 | 0.612 | 0.456 | 3.50E-82 |
| **GPR183** | 0 | 2.88587029 | 0.427 | 0.113 | 0 | | RGS2 | 1.10E-85 | 2.20841176 | 0.17 | 0.057 | 4.03E-81 |
| **CD83** | 0 | 3.14723159 | 0.376 | 0.09 | 0 | | SLC7A5 | 2.35E-85 | 1.79273174 | 0.284 | 0.131 | 8.61E-81 |
| **CD69** | 0 | 2.9688526 | 0.284 | 0.052 | 0 | | JUN | 1.59E-83 | 2.03690474 | 0.24 | 0.102 | 5.83E-79 |
| **HIST1H2BG** | 0 | 3.19120274 | 0.267 | 0.046 | 0 | | HSPE1 | 2.47E-83 | 1.87514775 | 0.332 | 0.173 | 9.05E-79 |
| **HIST1H2BJ** | 0 | 3.95196283 | 0.198 | 0.021 | 0 | | HSPB1 | 1.16E-82 | 1.96311652 | 0.346 | 0.185 | 4.24E-78 |
| HSPA1A | 9.35E-303 | 1.87637009 | 0.7 | 0.339 | 3.42E-298 | | NR4A1 | 6.26E-80 | 2.63842653 | 0.104 | 0.027 | 2.29E-75 |
| HIF1A-AS3 | 2.77E-283 | 2.42376659 | 0.546 | 0.223 | 1.01E-278 | | CCL3 | 2.77E-79 | 2.31195132 | 0.073 | 0.015 | 1.01E-74 |
| HSPA1B | 2.07E-229 | 1.83747575 | 0.489 | 0.194 | 7.58E-225 | | SSH2 | 1.03E-77 | 0.56512851 | 0.945 | 0.911 | 3.77E-73 |
| AC084871.1 | 9.12E-227 | 2.51532738 | 0.322 | 0.094 | 3.34E-222 | | TULP2 | 1.13E-77 | 3.96859184 | 0.031 | 0.003 | 4.13E-73 |
| **HIST1H2BC** | 2.82E-202 | 2.87067918 | 0.167 | 0.032 | 1.03E-197 | | ABHD15-AS1 | 2.38E-77 | 0.97466684 | 0.631 | 0.462 | 8.70E-73 |
| SPP1 | 1.27E-193 | 1.37929102 | 0.842 | 0.667 | 4.64E-189 | | HIST1H1C | 5.39E-76 | 2.04215302 | 0.113 | 0.031 | 1.97E-71 |
| HIF1A | 4.04E-193 | 1.67836318 | 0.766 | 0.55 | 1.48E-188 | | AC083862.3 | 1.26E-75 | 3.10760576 | 0.087 | 0.021 | 4.61E-71 |
| HSP90AA1 | 3.96E-190 | 2.4108932 | 0.777 | 0.612 | 1.45E-185 | | GNA13 | 6.84E-69 | 1.40638504 | 0.481 | 0.33 | 2.50E-64 |
| **HIST1H2BD** | 3.18E-185 | 2.36476941 | 0.248 | 0.068 | 1.16E-180 | | REL | 5.88E-68 | 1.20758613 | 0.497 | 0.352 | 2.15E-63 |
| RGS16 | 1.52E-169 | 2.77971664 | 0.2 | 0.05 | 5.56E-165 | | CTSB | 2.16E-65 | 0.88474951 | 0.738 | 0.662 | 7.89E-61 |
| AC022217.3 | 3.23E-168 | 3.56087137 | 0.085 | 0.011 | 1.18E-163 | | ACSL1 | 2.30E-63 | 0.98906766 | 0.595 | 0.453 | 8.43E-59 |
| TEX14 | 6.77E-167 | 3.43401834 | 0.151 | 0.031 | 2.48E-162 | | OTUD1 | 8.07E-63 | 1.99359563 | 0.146 | 0.054 | 2.95E-58 |
| MALAT1 | 3.52E-164 | 0.44339984 | 1 | 1 | 1.29E-159 | | HSPH1 | 2.19E-62 | 2.05625304 | 0.447 | 0.31 | 8.00E-58 |
| P4HA1 | 2.28E-161 | 1.75809061 | 0.645 | 0.41 | 8.35E-157 | | NFKBID | 5.04E-62 | 1.89096476 | 0.163 | 0.063 | 1.84E-57 |
| HIST1H1D | 3.75E-159 | 3.08367128 | 0.123 | 0.022 | 1.37E-154 | | NAMPT | 2.09E-61 | 1.56401476 | 0.345 | 0.198 | 7.63E-57 |
| PPP1R15A | 1.77E-145 | 2.62767949 | 0.161 | 0.038 | 6.48E-141 | | NR4A2 | 5.26E-61 | 2.11401875 | 0.139 | 0.05 | 1.93E-56 |
| CERNA2 | 2.35E-145 | 2.78730735 | 0.214 | 0.061 | 8.60E-141 | | HERPUD1 | 5.40E-61 | 1.2186093 | 0.461 | 0.317 | 1.98E-56 |
| OLR1 | 2.05E-144 | 1.72821384 | 0.511 | 0.275 | 7.52E-140 | | AL079338.1 | 1.44E-60 | 2.72029455 | 0.068 | 0.016 | 5.26E-56 |
| HIST2H2BE | 6.35E-136 | 2.17768196 | 0.203 | 0.058 | 2.32E-131 | | HIST1H2BL | 5.20E-58 | 3.86179209 | 0.016 | 0.001 | 1.90E-53 |
| ATF3 | 4.76E-130 | 3.19567153 | 0.091 | 0.015 | 1.74E-125 | | HIST1H2BK | 1.23E-57 | 3.43797096 | 0.026 | 0.003 | 4.51E-53 |
| GRID2 | 3.05E-126 | 1.04089632 | 0.708 | 0.455 | 1.12E-121 | | AC103591.3 | 1.75E-57 | 1.92554427 | 0.112 | 0.037 | 6.42E-53 |
| **BCAS2** | 2.35E-120 | 2.89908006 | 0.226 | 0.077 | 8.61E-116 | | VMP1 | 1.90E-54 | 0.85328068 | 0.665 | 0.561 | 6.95E-50 |
| HIST3H2BB | 1.12E-119 | 3.41597474 | 0.061 | 0.008 | 4.09E-115 | | CLEC2B | 9.92E-52 | 1.68711239 | 0.196 | 0.092 | 3.63E-47 |
| HIST2H2AA4 | 1.41E-118 | 2.36560125 | 0.126 | 0.028 | 5.15E-114 | | HIST1H3H | 1.97E-51 | 2.82529246 | 0.031 | 0.005 | 7.21E-47 |
| HIST1H1B | 2.14E-117 | 4.52508135 | 0.026 | 0.001 | 7.84E-113 | | ADM | 3.34E-50 | 2.83260993 | 0.036 | 0.006 | 1.22E-45 |
| DNAJB1 | 3.46E-107 | 2.24912903 | 0.243 | 0.092 | 1.27E-102 | | HIST2H2BF | 3.07E-49 | 1.08496532 | 0.337 | 0.202 | 1.13E-44 |
| HIST1H2AE | 3.09E-106 | 2.66157555 | 0.082 | 0.014 | 1.13E-101 | | ZFP36 | 2.31E-48 | 1.16676069 | 0.293 | 0.167 | 8.46E-44 |
| PFKFB3 | 1.43E-105 | 1.28173401 | 0.606 | 0.422 | 5.24E-101 | | BTG2 | 1.14E-47 | 1.45530356 | 0.257 | 0.142 | 4.18E-43 |
| PADI2 | 9.70E-104 | 1.51237413 | 0.551 | 0.356 | 3.55E-99 | | HIST1H2BM | 1.04E-46 | 3.32583969 | 0.019 | 0.002 | 3.82E-42 |
| UBC | 6.50E-103 | 1.42655021 | 0.593 | 0.422 | 2.38E-98 | | ZFP36L1 | 3.85E-45 | 0.66586419 | 0.728 | 0.648 | 1.41E-40 |

| | | | | | | | | | | | | |
|---|---|---|---|---|---|---|---|---|---|---|---|---|
| PTGS2 | 1.04E-44 | 3.1500031 | 0.043 | 0.009 | 3.82E-40 | | CCL4 | 1.55E-27 | 2.24231376 | 0.037 | 0.01 | 5.66E-23 |
| TFRC | 1.05E-43 | 2.17484127 | 0.252 | 0.145 | 3.85E-39 | | AC011933.4 | 5.46E-27 | 0.70967914 | 0.421 | 0.32 | 2.00E-22 |
| VEGFA | 1.94E-43 | 1.53026064 | 0.15 | 0.066 | 7.09E-39 | | FCGR2A | 5.47E-27 | 0.92869837 | 0.404 | 0.316 | 2.00E-22 |
| H3F3B | 1.06E-42 | 1.50002056 | 0.414 | 0.303 | 3.88E-38 | | HIST1H2BH | 1.33E-26 | 2.02387253 | 0.053 | 0.018 | 4.88E-22 |
| HSPD1 | 7.60E-41 | 1.72760506 | 0.319 | 0.208 | 2.78E-36 | | ADGRB3 | 4.86E-26 | 0.56397691 | 0.618 | 0.542 | 1.78E-21 |
| BCL2A1 | 1.03E-40 | 1.54935335 | 0.103 | 0.039 | 3.77E-36 | | FAM13A | 9.85E-26 | 0.85648406 | 0.609 | 0.556 | 3.60E-21 |
| SAT1 | 1.06E-40 | 0.79093686 | 0.785 | 0.756 | 3.87E-36 | | RANBP2 | 1.15E-25 | 1.07386058 | 0.436 | 0.361 | 4.21E-21 |
| PDGFB | 5.16E-40 | 1.17471759 | 0.362 | 0.246 | 1.89E-35 | | ARHGAP26-AS1 | 1.45E-25 | 1.03361771 | 0.161 | 0.089 | 5.30E-21 |
| AC120193.1 | 7.02E-39 | 0.67138695 | 0.619 | 0.511 | 2.57E-34 | | IRAK2 | 1.69E-25 | 1.30667099 | 0.164 | 0.093 | 6.19E-21 |
| HK2 | 1.13E-38 | 1.27507066 | 0.292 | 0.181 | 4.15E-34 | | HIST1H3D | 2.65E-25 | 2.48905574 | 0.026 | 0.006 | 9.70E-21 |
| MCF2L2 | 6.75E-38 | 1.17344022 | 0.441 | 0.329 | 2.47E-33 | | DENND3 | 2.22E-24 | 0.4110243 | 0.738 | 0.685 | 8.14E-20 |
| IER5 | 8.75E-38 | 1.76258123 | 0.165 | 0.081 | 3.20E-33 | | AC037487.2 | 4.19E-24 | 2.36487647 | 0.027 | 0.007 | 1.53E-19 |
| RHOB | 3.72E-37 | 1.17355513 | 0.266 | 0.161 | 1.36E-32 | | PGK1 | 7.06E-24 | 0.84729266 | 0.444 | 0.369 | 2.58E-19 |
| ELL2 | 6.27E-37 | 0.83168148 | 0.657 | 0.572 | 2.30E-32 | | AC239799.2 | 1.94E-23 | 3.22844902 | 0.013 | 0.002 | 7.10E-19 |
| HMOX1 | 5.88E-36 | 1.8178411 | 0.206 | 0.116 | 2.15E-31 | | CKS2 | 3.10E-23 | 2.18939876 | 0.053 | 0.019 | 1.13E-18 |
| PER1 | 2.11E-35 | 1.1902557 | 0.254 | 0.153 | 7.74E-31 | | AL021155.5 | 5.54E-23 | 1.83154253 | 0.035 | 0.01 | 2.03E-18 |
| HIST1H2AG | 2.19E-35 | 2.60332174 | 0.028 | 0.005 | 8.01E-31 | | SERPINE1 | 1.59E-22 | 2.50150602 | 0.091 | 0.043 | 5.83E-18 |
| CEBPD | 4.03E-35 | 0.86240558 | 0.368 | 0.251 | 1.47E-30 | | AL390957.1 | 3.43E-22 | 1.34771715 | 0.104 | 0.051 | 1.26E-17 |
| HIST1H2AH | 5.52E-35 | 3.29132482 | 0.018 | 0.002 | 2.02E-30 | | TNFRSF1B | 3.70E-22 | 0.748706 | 0.46 | 0.388 | 1.35E-17 |
| HIST1H4E | 6.06E-35 | 2.04638125 | 0.061 | 0.019 | 2.22E-30 | | RAPGEF1 | 3.99E-22 | 0.52068076 | 0.67 | 0.627 | 1.46E-17 |
| BHLHE41 | 8.13E-35 | 0.63118821 | 0.652 | 0.564 | 2.98E-30 | | AL731563.2 | 4.14E-22 | 2.92607242 | 0.013 | 0.002 | 1.52E-17 |
| SLC2A5 | 8.62E-35 | 0.83137039 | 0.585 | 0.482 | 3.16E-30 | | APMAP | 9.45E-22 | 0.70833442 | 0.394 | 0.306 | 3.46E-17 |
| SGK1 | 1.15E-34 | 1.16413044 | 0.417 | 0.312 | 4.22E-30 | | EGR1 | 1.33E-21 | 1.71470969 | 0.079 | 0.035 | 4.88E-17 |
| TRA2B | 1.40E-34 | 1.20936403 | 0.472 | 0.383 | 5.12E-30 | | RLF | 2.60E-21 | 0.63790671 | 0.431 | 0.352 | 9.51E-17 |
| KDM4B | 3.12E-34 | 0.88788106 | 0.37 | 0.261 | 1.14E-29 | | GRASP | 3.39E-21 | 1.39017308 | 0.098 | 0.048 | 1.24E-16 |
| HSP90AB1 | 5.39E-34 | 0.77354026 | 0.582 | 0.495 | 1.97E-29 | | NEAT1 | 7.19E-21 | 0.26059402 | 0.976 | 0.965 | 2.63E-16 |
| TBC1D8 | 6.29E-34 | 0.79906961 | 0.391 | 0.276 | 2.30E-29 | | BCL6 | 7.22E-21 | 0.87146422 | 0.296 | 0.214 | 2.64E-16 |
| HAMP | 3.02E-33 | 0.96761016 | 0.318 | 0.209 | 1.10E-28 | | CCL3L1 | 8.23E-21 | 2.55107484 | 0.021 | 0.005 | 3.01E-16 |
| FTH1 | 3.46E-33 | 0.17368639 | 0.83 | 0.773 | 1.27E-28 | | BAG3 | 1.63E-20 | 2.25195413 | 0.095 | 0.047 | 5.95E-16 |
| HIST1H4J | 4.70E-31 | 2.89584068 | 0.021 | 0.003 | 1.72E-26 | | ETS2 | 3.87E-20 | 0.74824736 | 0.442 | 0.373 | 1.42E-15 |
| NR4A3 | 2.01E-30 | 2.20654725 | 0.054 | 0.017 | 7.34E-26 | | HIST3H2A | 5.06E-20 | 1.78346495 | 0.064 | 0.027 | 1.85E-15 |
| JUNB | 2.81E-30 | 1.17125383 | 0.198 | 0.113 | 1.03E-25 | | SMAP2 | 1.33E-19 | 0.37798499 | 0.731 | 0.693 | 4.86E-15 |
| AC092053.3 | 3.18E-30 | 2.75976557 | 0.02 | 0.003 | 1.16E-25 | | FOSB | 1.73E-19 | 2.13843721 | 0.04 | 0.014 | 6.32E-15 |
| CXCR4 | 3.29E-30 | 1.4581055 | 0.103 | 0.044 | 1.20E-25 | | HSPA6 | 2.46E-19 | 1.61667104 | 0.106 | 0.056 | 9.01E-15 |
| SCIN | 3.83E-30 | 0.71905916 | 0.509 | 0.403 | 1.40E-25 | | TNF | 5.85E-19 | 1.99163006 | 0.036 | 0.012 | 2.14E-14 |
| HIST2H2AC | 4.29E-30 | 1.98701066 | 0.054 | 0.017 | 1.57E-25 | | NFIL3 | 9.51E-19 | 1.4155524 | 0.097 | 0.05 | 3.48E-14 |
| TMEM163 | 4.61E-30 | 0.59447806 | 0.548 | 0.423 | 1.69E-25 | | JDP2 | 9.70E-19 | 1.02043723 | 0.326 | 0.252 | 3.55E-14 |
| SPON1 | 6.08E-30 | 1.51974492 | 0.124 | 0.059 | 2.23E-25 | | HIST1H1E | 1.10E-18 | 1.46711826 | 0.049 | 0.019 | 4.01E-14 |
| GLA | 1.58E-29 | 1.80872809 | 0.081 | 0.032 | 5.77E-25 | | HLA-DQA1 | 1.24E-18 | 1.24635805 | 0.137 | 0.08 | 4.52E-14 |
| PRDM1 | 2.20E-29 | 1.21780632 | 0.192 | 0.109 | 8.04E-25 | | CCL4L2 | 1.57E-18 | 2.40069226 | 0.025 | 0.007 | 5.74E-14 |
| AC245014.3 | 2.40E-29 | 2.19865828 | 0.034 | 0.008 | 8.78E-25 | | HIST1H2AI | 1.64E-18 | 2.74942604 | 0.012 | 0.002 | 6.02E-14 |
| AC253572.2 | 5.12E-29 | 2.33204614 | 0.031 | 0.007 | 1.87E-24 | | LRRC4C | 5.88E-18 | 1.17536597 | 0.299 | 0.226 | 2.15E-13 |
| AC090579.1 | 1.49E-28 | 0.88613323 | 0.359 | 0.255 | 5.45E-24 | | LINC02742 | 3.80E-17 | 0.7189836 | 0.211 | 0.14 | 1.39E-12 |
| AP000331.1 | 5.41E-28 | 0.89931464 | 0.365 | 0.267 | 1.98E-23 | | INSIG1 | 3.96E-17 | 1.70294908 | 0.102 | 0.056 | 1.45E-12 |
| CXCL8 | 5.63E-28 | 3.08576477 | 0.013 | 0.002 | 2.06E-23 | | AL691403.1 | 9.33E-17 | 2.42414343 | 0.014 | 0.003 | 3.42E-12 |
| ID2 | 6.40E-28 | 1.26564931 | 0.324 | 0.231 | 2.34E-23 | | ABCC4 | 1.34E-16 | 0.36909133 | 0.733 | 0.683 | 4.90E-12 |
| PAPOLG | 1.02E-27 | 1.129078 | 0.281 | 0.189 | 3.74E-23 | | CYCS | 1.81E-16 | 1.05403761 | 0.236 | 0.172 | 6.61E-12 |

| | | | | | | | | | | | | |
|---|---|---|---|---|---|---|---|---|---|---|---|---|
| GRB2 | 2.28E-16 | 0.45701301 | 0.643 | 0.614 | 8.35E-12 | | FAM162A | 6.36E-12 | 0.96777226 | 0.154 | 0.105 | 2.33E-07 |
| C5AR1 | 2.53E-16 | 1.05629733 | 0.157 | 0.099 | 9.24E-12 | | SH3TC1 | 6.50E-12 | 0.43693337 | 0.467 | 0.422 | 2.38E-07 |
| PDK3 | 3.56E-16 | 0.69495582 | 0.342 | 0.271 | 1.30E-11 | | PDK4 | 7.04E-12 | 1.66802741 | 0.231 | 0.182 | 2.58E-07 |
| RAB20 | 4.20E-16 | 0.95663573 | 0.223 | 0.159 | 1.54E-11 | | RNASET2 | 8.29E-12 | 0.2585743 | 0.756 | 0.74 | 3.03E-07 |
| RHBDF2 | 6.21E-16 | 0.37983491 | 0.676 | 0.635 | 2.27E-11 | | RAB11A | 8.38E-12 | 0.43233067 | 0.505 | 0.456 | 3.07E-07 |
| RNF122 | 6.22E-16 | 1.36002042 | 0.068 | 0.032 | 2.28E-11 | | AC104365.1 | 9.00E-12 | 1.06480897 | 0.105 | 0.065 | 3.29E-07 |
| HEATR5B | 8.30E-16 | 0.55690411 | 0.483 | 0.419 | 3.04E-11 | | AL513327.1 | 1.26E-11 | 0.56997738 | 0.292 | 0.234 | 4.61E-07 |
| IER2 | 9.97E-16 | 1.04494857 | 0.145 | 0.091 | 3.65E-11 | | EGR2 | 1.29E-11 | 2.16282361 | 0.016 | 0.004 | 4.71E-07 |
| TMEM52B | 1.15E-15 | 1.38322015 | 0.047 | 0.02 | 4.21E-11 | | AL049651.1 | 1.34E-11 | 2.20297611 | 0.013 | 0.003 | 4.89E-07 |
| ADAM28 | 1.77E-15 | 0.35103147 | 0.822 | 0.791 | 6.49E-11 | | ALOX15B | 1.38E-11 | 0.95798866 | 0.093 | 0.055 | 5.05E-07 |
| CSF2RA | 2.01E-15 | 0.5121745 | 0.497 | 0.432 | 7.36E-11 | | CHSY1 | 2.60E-11 | 0.60648262 | 0.434 | 0.393 | 9.51E-07 |
| SLC11A1 | 2.04E-15 | 0.43677079 | 0.54 | 0.48 | 7.45E-11 | | YEATS2 | 2.94E-11 | 0.62866391 | 0.304 | 0.249 | 1.08E-06 |
| QKI | 2.31E-15 | 0.24981207 | 0.937 | 0.94 | 8.47E-11 | | IER3 | 3.12E-11 | 0.8452336 | 0.169 | 0.12 | 1.14E-06 |
| AC002451.1 | 2.39E-15 | 1.64549931 | 0.071 | 0.035 | 8.75E-11 | | AC008892.1 | 3.52E-11 | 0.77880421 | 0.184 | 0.133 | 1.29E-06 |
| PLIN2 | 3.42E-15 | 1.84212563 | 0.071 | 0.035 | 1.25E-10 | | MAFF | 4.95E-11 | 1.47263928 | 0.027 | 0.01 | 1.81E-06 |
| RAB1A | 4.08E-15 | 0.40675425 | 0.596 | 0.551 | 1.49E-10 | | DENND5A | 7.52E-11 | 0.47652173 | 0.438 | 0.389 | 2.75E-06 |
| FOXP2 | 4.10E-15 | 0.30796054 | 0.71 | 0.636 | 1.50E-10 | | MT-CO1 | 9.32E-11 | 0.11283187 | 0.883 | 0.84 | 3.41E-06 |
| SYAP1 | 4.12E-15 | 1.30530042 | 0.156 | 0.101 | 1.51E-10 | | RIPK2 | 9.52E-11 | 1.05823586 | 0.156 | 0.111 | 3.48E-06 |
| CLK1 | 4.31E-15 | 0.76128109 | 0.405 | 0.351 | 1.58E-10 | | FLT1 | 9.76E-11 | 1.11815335 | 0.134 | 0.09 | 3.57E-06 |
| SNAP25 | 5.90E-15 | 0.2865883 | 0.789 | 0.745 | 2.16E-10 | | EGR3 | 1.31E-10 | 1.24052429 | 0.072 | 0.041 | 4.80E-06 |
| HLA-DQB1 | 6.23E-15 | 0.895078 | 0.191 | 0.132 | 2.28E-10 | | TMEM140 | 1.67E-10 | 1.1663645 | 0.088 | 0.053 | 6.10E-06 |
| FAM110B | 1.69E-14 | 0.62115065 | 0.288 | 0.218 | 6.19E-10 | | ANKRD37 | 1.73E-10 | 1.65506399 | 0.06 | 0.033 | 6.32E-06 |
| EIF4A3 | 1.86E-14 | 1.74942575 | 0.093 | 0.052 | 6.80E-10 | | KLHL6-AS1 | 2.19E-10 | 1.67619506 | 0.035 | 0.015 | 8.01E-06 |
| HIST2H2AB | 2.01E-14 | 2.33622796 | 0.012 | 0.002 | 7.35E-10 | | KLF2 | 2.28E-10 | 1.19253811 | 0.049 | 0.024 | 8.34E-06 |
| USP36 | 2.44E-14 | 1.21933176 | 0.191 | 0.135 | 8.93E-10 | | MT-ND1 | 2.30E-10 | 0.10814896 | 0.867 | 0.824 | 8.44E-06 |
| PPP1CB | 2.63E-14 | 0.51882874 | 0.457 | 0.397 | 9.64E-10 | | PICALM | 2.53E-10 | 0.22059599 | 0.901 | 0.892 | 9.25E-06 |
| PPP1R3C | 3.24E-14 | 1.92693576 | 0.052 | 0.024 | 1.19E-09 | | BASP1 | 2.55E-10 | 0.33176213 | 0.582 | 0.548 | 9.32E-06 |
| GBP2 | 4.24E-14 | 1.1153462 | 0.121 | 0.073 | 1.55E-09 | | AP000919.2 | 2.62E-10 | 0.90573358 | 0.121 | 0.079 | 9.58E-06 |
| SLC2A1 | 7.75E-14 | 1.4035841 | 0.049 | 0.022 | 2.84E-09 | | MIR210HG | 3.60E-10 | 1.21155636 | 0.035 | 0.016 | 1.32E-05 |
| HSPA8 | 8.39E-14 | 0.6992336 | 0.366 | 0.307 | 3.07E-09 | | PRKCA-AS1 | 3.60E-10 | 0.63388461 | 0.17 | 0.122 | 1.32E-05 |
| ARHGEF7 | 1.27E-13 | 0.5384727 | 0.465 | 0.411 | 4.64E-09 | | OXR1 | 4.42E-10 | 0.2080895 | 0.813 | 0.793 | 1.62E-05 |
| AL606923.2 | 1.39E-13 | 1.92696634 | 0.02 | 0.006 | 5.10E-09 | | GABARAPL1 | 5.10E-10 | 0.54326179 | 0.26 | 0.207 | 1.87E-05 |
| RBPJ | 2.05E-13 | 0.36756536 | 0.55 | 0.497 | 7.50E-09 | | UCHL1 | 5.80E-10 | 0.28551966 | 0.634 | 0.601 | 2.12E-05 |
| CEMIP2 | 3.71E-13 | 1.01106561 | 0.209 | 0.153 | 1.36E-08 | | IFRD1 | 9.01E-10 | 1.49498186 | 0.205 | 0.161 | 3.30E-05 |
| WSB1 | 3.90E-13 | 0.40729032 | 0.665 | 0.641 | 1.43E-08 | | AC012447.1 | 9.70E-10 | 1.19234516 | 0.06 | 0.033 | 3.55E-05 |
| SRGAP2-AS1 | 4.72E-13 | 0.68370697 | 0.245 | 0.183 | 1.73E-08 | | LDHA | 9.74E-10 | 0.6527378 | 0.198 | 0.148 | 3.56E-05 |
| CH25H | 5.39E-13 | 1.12048094 | 0.082 | 0.045 | 1.97E-08 | | MT-ND2 | 1.19E-09 | 0.12455464 | 0.871 | 0.829 | 4.36E-05 |
| RASGEF1C | 6.70E-13 | 0.33517732 | 0.498 | 0.424 | 2.45E-08 | | AC012181.1 | 1.20E-09 | 1.5591557 | 0.028 | 0.012 | 4.38E-05 |
| KDM3A | 7.65E-13 | 0.75935725 | 0.236 | 0.178 | 2.80E-08 | | AC011257.1 | 1.42E-09 | 1.18662928 | 0.045 | 0.023 | 5.18E-05 |
| CLEC7A | 8.37E-13 | 0.53791647 | 0.426 | 0.37 | 3.06E-08 | | MCL1 | 1.43E-09 | 0.78024355 | 0.191 | 0.143 | 5.23E-05 |
| CCNY | 1.18E-12 | 0.36503216 | 0.659 | 0.644 | 4.30E-08 | | RTN1 | 1.94E-09 | 0.3740882 | 0.503 | 0.462 | 7.08E-05 |
| SIPA1L1 | 1.46E-12 | 0.37287794 | 0.617 | 0.574 | 5.36E-08 | | ARHGAP26 | 2.08E-09 | 0.18737758 | 0.926 | 0.919 | 7.62E-05 |
| CCDC26 | 1.88E-12 | 0.37218104 | 0.368 | 0.289 | 6.90E-08 | | SNAPC1 | 2.21E-09 | 0.99895993 | 0.091 | 0.057 | 8.09E-05 |
| TBC1D22A | 2.07E-12 | 0.29276345 | 0.78 | 0.76 | 7.57E-08 | | FOXP1 | 3.32E-09 | 0.25106583 | 0.693 | 0.647 | 0.00012168 |
| AL360012.1 | 2.82E-12 | 1.92253121 | 0.017 | 0.005 | 1.03E-07 | | LPCAT1 | 3.96E-09 | 0.80465852 | 0.156 | 0.114 | 0.00014487 |
| B3GNT5 | 2.92E-12 | 0.8216863 | 0.394 | 0.343 | 1.07E-07 | | MT-CO2 | 4.33E-09 | 0.10370058 | 0.883 | 0.847 | 0.00015844 |
| AL499604.1 | 4.42E-12 | 2.78239204 | 0.015 | 0.004 | 1.62E-07 | | LINC02008 | 5.98E-09 | 0.88131268 | 0.066 | 0.039 | 0.00021905 |

| | | | | | | | | | | | | |
|---|---|---|---|---|---|---|---|---|---|---|---|---|
| TAGAP | 6.49E-09 | 1.1438063 | 0.085 | 0.054 | 0.00023758 | | H3F3A | 2.89E-07 | 0.4729849 | 0.357 | 0.32 | 0.010595 |
| IL3RA | 6.59E-09 | 0.9193325 | 0.122 | 0.085 | 0.00024115 | | MAP1B | 3.07E-07 | 0.22710664 | 0.589 | 0.553 | 0.01123882 |
| ARRDC3 | 6.70E-09 | 1.10712937 | 0.126 | 0.088 | 0.00024532 | | PTGES3 | 3.78E-07 | 0.50452098 | 0.403 | 0.368 | 0.01385059 |
| PEAK3 | 8.66E-09 | 1.75720805 | 0.023 | 0.009 | 0.0003171 | | CYTIP | 4.48E-07 | 0.77456589 | 0.127 | 0.093 | 0.01640023 |
| AC104695.4 | 9.31E-09 | 1.80168743 | 0.012 | 0.004 | 0.00034059 | | TTC7A | 4.51E-07 | 0.40183925 | 0.479 | 0.449 | 0.01649052 |
| CKB | 9.55E-09 | 0.27539244 | 0.561 | 0.527 | 0.00034971 | | PFKFB4 | 5.18E-07 | 1.11012746 | 0.068 | 0.043 | 0.01896938 |
| MB21D2 | 1.12E-08 | 0.61106084 | 0.2 | 0.153 | 0.00041037 | | ADSSL1 | 5.42E-07 | 1.43590452 | 0.022 | 0.01 | 0.01983006 |
| UST | 1.19E-08 | 0.46336529 | 0.345 | 0.297 | 0.00043379 | | OSM | 5.50E-07 | 1.60401329 | 0.018 | 0.007 | 0.0201259 |
| SYT1 | 1.53E-08 | 0.2221925 | 0.685 | 0.652 | 0.00056062 | | ZNF331 | 5.72E-07 | 1.46477685 | 0.182 | 0.149 | 0.0209469 |
| CHN1 | 1.68E-08 | 0.24409399 | 0.63 | 0.596 | 0.00061312 | | AP002336.2 | 5.76E-07 | 0.48099957 | 0.233 | 0.191 | 0.02108843 |
| VSNL1 | 2.01E-08 | 0.33247399 | 0.534 | 0.498 | 0.00073584 | | AC078883.1 | 5.77E-07 | 0.76452569 | 0.102 | 0.071 | 0.02111108 |
| LINC00910 | 2.03E-08 | 1.19145183 | 0.048 | 0.026 | 0.00074329 | | CALM3 | 5.77E-07 | 0.29174704 | 0.53 | 0.504 | 0.02112141 |
| SLC8A1-AS1 | 2.15E-08 | 0.47710399 | 0.336 | 0.292 | 0.00078521 | | GZF1 | 6.16E-07 | 0.86962966 | 0.058 | 0.035 | 0.02254149 |
| AC092120.3 | 2.20E-08 | 0.71428897 | 0.103 | 0.069 | 0.00080436 | | PLA2G6 | 6.25E-07 | 1.02488409 | 0.056 | 0.034 | 0.02287289 |
| AL731574.1 | 2.21E-08 | 0.96284011 | 0.084 | 0.053 | 0.00081059 | | NRARP | 6.77E-07 | 1.534339 | 0.023 | 0.01 | 0.02476464 |
| AC009542.1 | 2.28E-08 | 1.27610005 | 0.043 | 0.023 | 0.00083627 | | PDK1 | 6.80E-07 | 0.6533833 | 0.178 | 0.141 | 0.02488596 |
| ZNF292 | 2.29E-08 | 0.39303496 | 0.562 | 0.55 | 0.00083947 | | NCMAP | 7.14E-07 | 0.96068455 | 0.054 | 0.032 | 0.02612595 |
| MEG3 | 2.30E-08 | 0.25085006 | 0.634 | 0.596 | 0.00084264 | | MAP3K8 | 7.26E-07 | 0.55748514 | 0.335 | 0.299 | 0.02658597 |
| AL353759.1 | 3.13E-08 | 1.53744668 | 0.03 | 0.014 | 0.00114725 | | TBC1D14 | 7.39E-07 | 0.28401157 | 0.541 | 0.52 | 0.02702991 |
| SPARCL1 | 3.45E-08 | 0.26828313 | 0.637 | 0.608 | 0.00126442 | | AC079209.2 | 7.93E-07 | 1.07870049 | 0.049 | 0.029 | 0.02902775 |
| NPY1R | 4.38E-08 | 0.99372938 | 0.061 | 0.036 | 0.00160444 | | LINC01344 | 8.57E-07 | 1.98855542 | 0.011 | 0.004 | 0.03135892 |
| FGF11 | 4.55E-08 | 1.46472804 | 0.024 | 0.01 | 0.00166464 | | ROR2 | 9.38E-07 | 0.9921439 | 0.065 | 0.041 | 0.03434557 |
| ABR | 5.10E-08 | 0.19922913 | 0.806 | 0.799 | 0.00186711 | | LSAMP | 9.58E-07 | 0.19828942 | 0.518 | 0.481 | 0.03507538 |
| GEM | 5.96E-08 | 1.97902107 | 0.016 | 0.006 | 0.00218235 | | POU5F1 | 9.82E-07 | 1.25811134 | 0.024 | 0.011 | 0.03594137 |
| C3AR1 | 5.97E-08 | 0.81437777 | 0.221 | 0.181 | 0.00218395 | | PIK3IP1 | 9.89E-07 | 0.60696617 | 0.161 | 0.124 | 0.03618074 |
| Z93020.1 | 6.11E-08 | 1.37469819 | 0.025 | 0.011 | 0.00223464 | | SLC25A25 | 1.01E-06 | 0.81090596 | 0.133 | 0.1 | 0.03691372 |
| PMAIP1 | 6.30E-08 | 2.53239835 | 0.012 | 0.004 | 0.00230625 | | TPI1 | 1.07E-06 | 0.46444102 | 0.337 | 0.301 | 0.03918036 |
| ELMO1-AS1 | 6.41E-08 | 0.51854081 | 0.292 | 0.247 | 0.002345 | | COL27A1 | 1.12E-06 | 1.35611823 | 0.033 | 0.017 | 0.04081046 |
| RNMT | 6.97E-08 | 0.49360741 | 0.339 | 0.297 | 0.00255113 | | DNAJB6 | 1.20E-06 | 0.59386651 | 0.4 | 0.372 | 0.04406546 |
| B4GALT1 | 7.11E-08 | 0.63336535 | 0.357 | 0.32 | 0.0026025 | | SOCS3 | 1.22E-06 | 0.68021847 | 0.066 | 0.042 | 0.04477531 |
| ENO1 | 7.26E-08 | 0.50807781 | 0.31 | 0.269 | 0.00265602 | | HLA-DRB1 | 1.23E-06 | 0.41496504 | 0.363 | 0.331 | 0.0451402 |
| AL139807.1 | 7.38E-08 | 0.76099546 | 0.093 | 0.061 | 0.00270228 | | ATP1B1 | 1.26E-06 | 0.32760866 | 0.429 | 0.393 | 0.04626698 |
| BTG1 | 8.87E-08 | 0.60368083 | 0.213 | 0.169 | 0.00324829 | | ZFAND2A | 1.31E-06 | 1.403179 | 0.059 | 0.037 | 0.04808226 |
| DNAJB4 | 9.42E-08 | 1.14950185 | 0.09 | 0.06 | 0.00344613 | | MT3 | 1.35E-06 | 0.24506284 | 0.593 | 0.566 | 0.04926094 |
| IRF8 | 1.24E-07 | 0.61008831 | 0.312 | 0.272 | 0.00455367 | | | | | | | |
| AC023051.1 | 1.61E-07 | 0.4292696 | 0.265 | 0.218 | 0.00587977 | | | | | | | |
| AL158832.1 | 1.68E-07 | 0.79079453 | 0.084 | 0.055 | 0.0061427 | | | | | | | |
| TPM4 | 1.77E-07 | 0.65951477 | 0.191 | 0.151 | 0.00647771 | | | | | | | |
| AC073626.1 | 1.84E-07 | 1.93689249 | 0.01 | 0.003 | 0.00672033 | | | | | | | |
| THEMIS2 | 1.85E-07 | 0.4788976 | 0.197 | 0.154 | 0.00676273 | | | | | | | |
| AL136987.1 | 2.13E-07 | 1.78776587 | 0.015 | 0.005 | 0.00780275 | | | | | | | |
| ANK3 | 2.15E-07 | 0.44305748 | 0.332 | 0.286 | 0.00785674 | | | | | | | |
| KLF6 | 2.44E-07 | 0.65700606 | 0.294 | 0.256 | 0.00893253 | | | | | | | |
| KCNMB1 | 2.52E-07 | 0.47000186 | 0.23 | 0.186 | 0.00920541 | | | | | | | |
| IL1B | 2.62E-07 | 0.82450992 | 0.088 | 0.059 | 0.00958332 | | | | | | | |
| PRKCA | 2.82E-07 | 0.24762456 | 0.7 | 0.675 | 0.01030369 | | | | | | | |
| NRG3 | 2.82E-07 | 0.26429367 | 0.48 | 0.443 | 0.01031502 | | | | | | | |
| ZBTB16 | 2.87E-07 | 0.35110054 | 0.483 | 0.445 | 0.01050876 | | | | | | | |

**eFigure 1.** Quantification of β-amyloid plaque burden, neuronal density, and GFAP-positive astrocyte burden in the STS

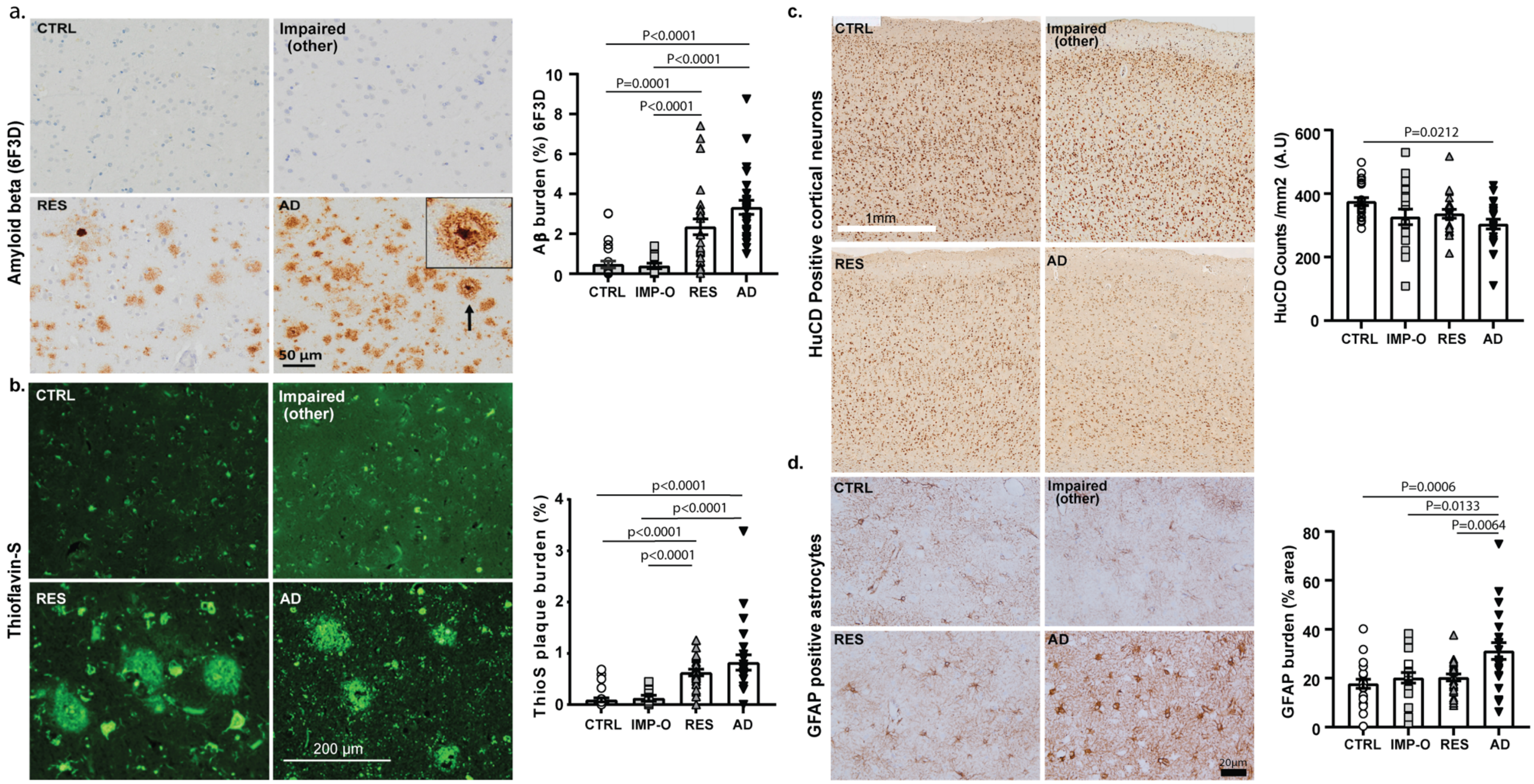


Representative photomicrographs and quantification of (a) 6F3D and (b) Thioflavin-S positive amyloid plaque burdens in the STS. No significant differences were detected in those measures between resilient and demented AD cases. Representative photomicrographs and quantification of (c) number of HuCD-positive neurons, and (d) GFAP-positive astrocyte burden in the STS. Demented AD showed a significantly higher GFAP-positive astrocyte burden compared to resilient cases.

**eFigure 2.** Tau seeding assays and analyses examining relationships with NFT counts and total pTau burden in the STS

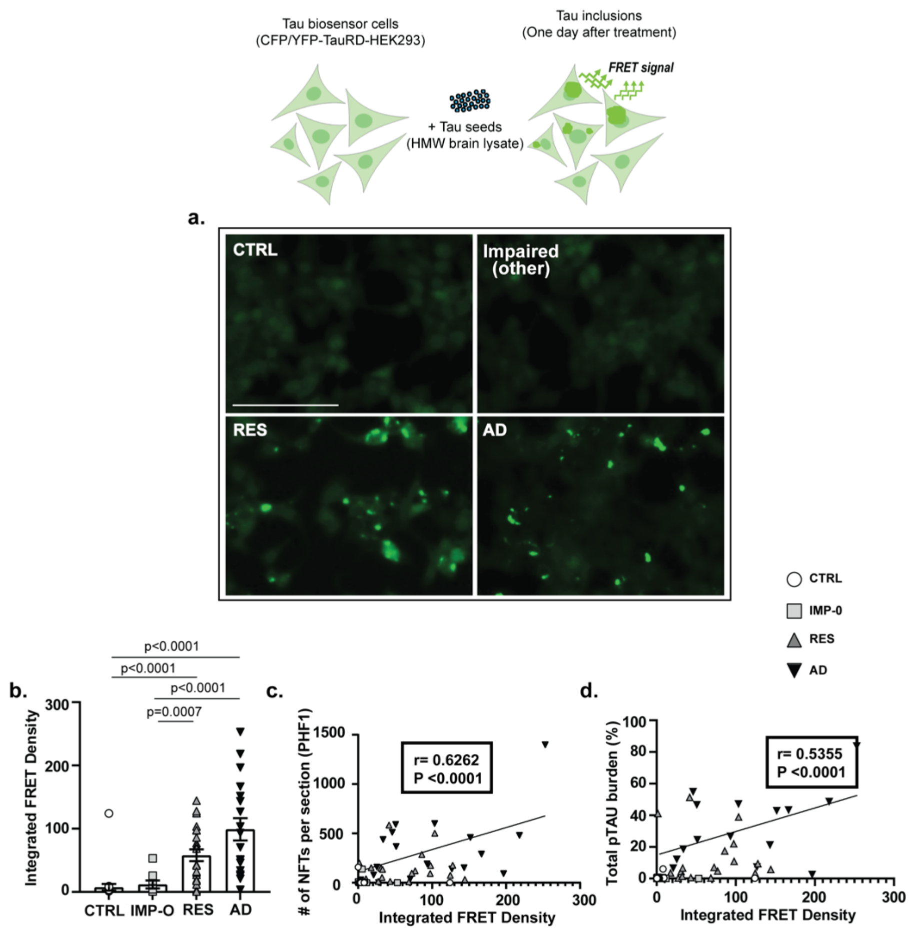


Representative images (a) and (b) quantification of tau aggregates in HEK293 cells exposed to STS brain lysates. No aggregates were detected in cells exposed to STS brain lysates from control or IMP-O brains. A significantly higher amount of tau aggregates was detected in cells exposed to STS brain lysates from demented AD and resilient brains without significant differences between these two groups. Tau seeding integrated FRET density was significantly correlated with (c) number of NFTs, and (d) total pTau burden.

**eFigure 3.** Correlation analyses of synapse density with total pTau burden, synaptic pTau oligomer levels, and CD68+ microglia burdens in the STS

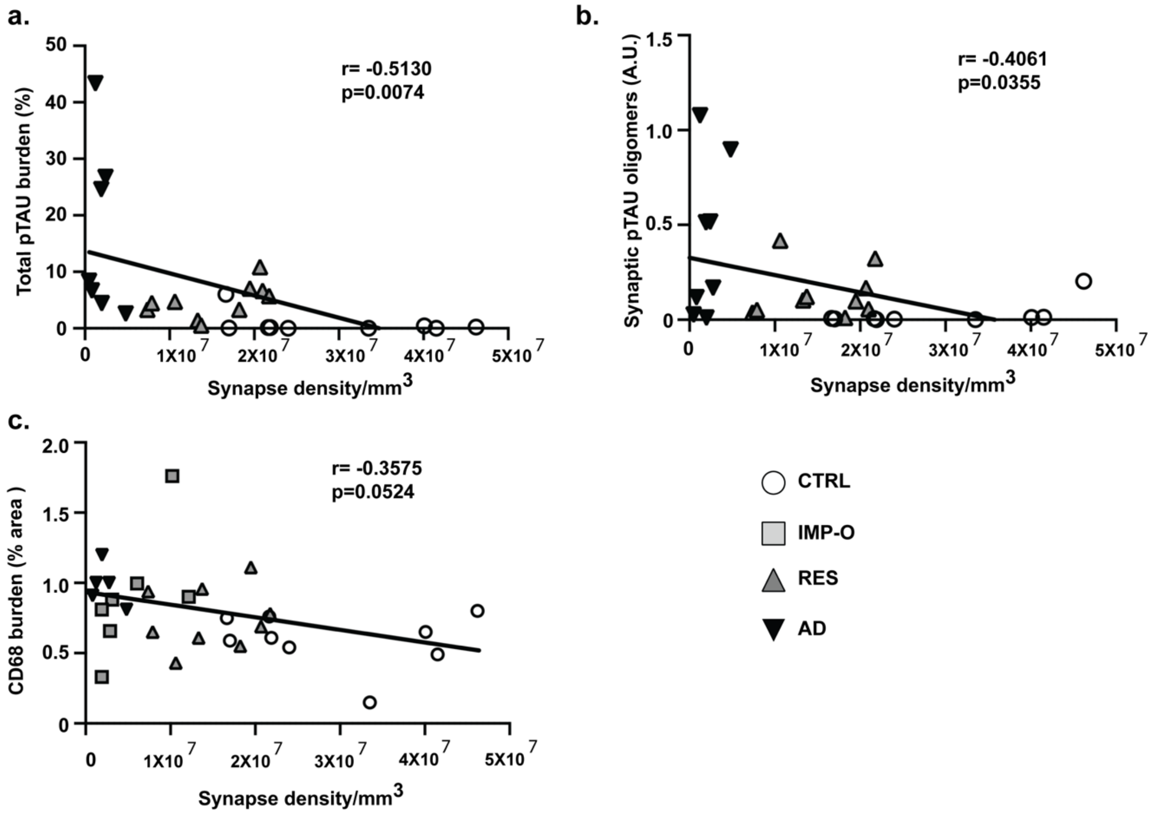


Correlation analyses revealed significant correlations between synapse density and (a) total pTAU burden, (b) synapse oligomeric pTAU (PHF1), and (c) CD68+ microglia burden in the STS.

**eFigure 4.** Genes implicated in the regulation of altered biological processes in demented AD compared to resilient brains

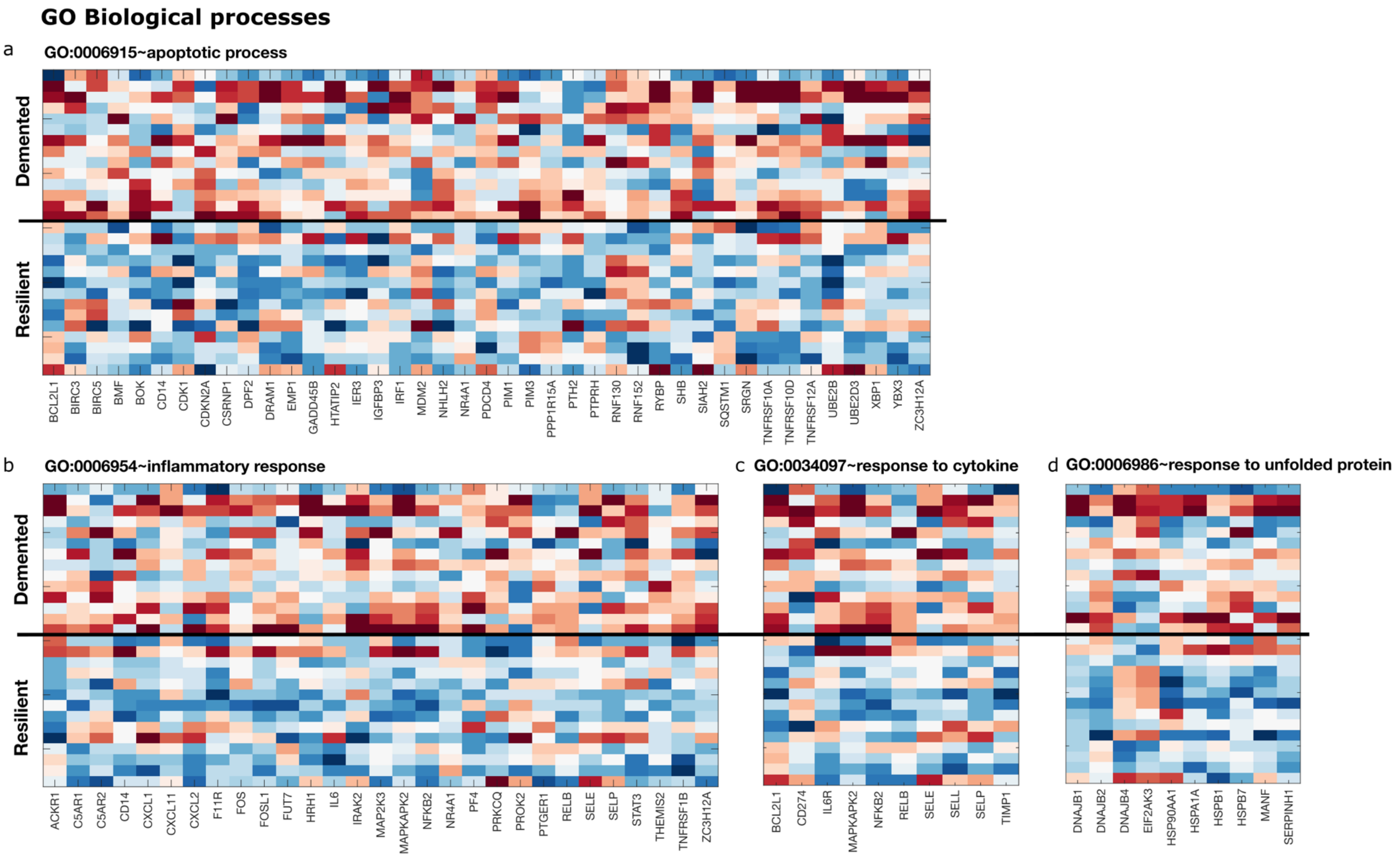


Representative heatmap shows genes with significant downregulation in resilient compared to demented AD and were linked to Gene Ontology biological processes (a) apoptotic pathway (b) Inflammatory response (c) response to cytokine and (d) response to unfolded protein

**eFigure 5**. Genes implicated in dysregulated pathways in demented AD compared to resilient brains

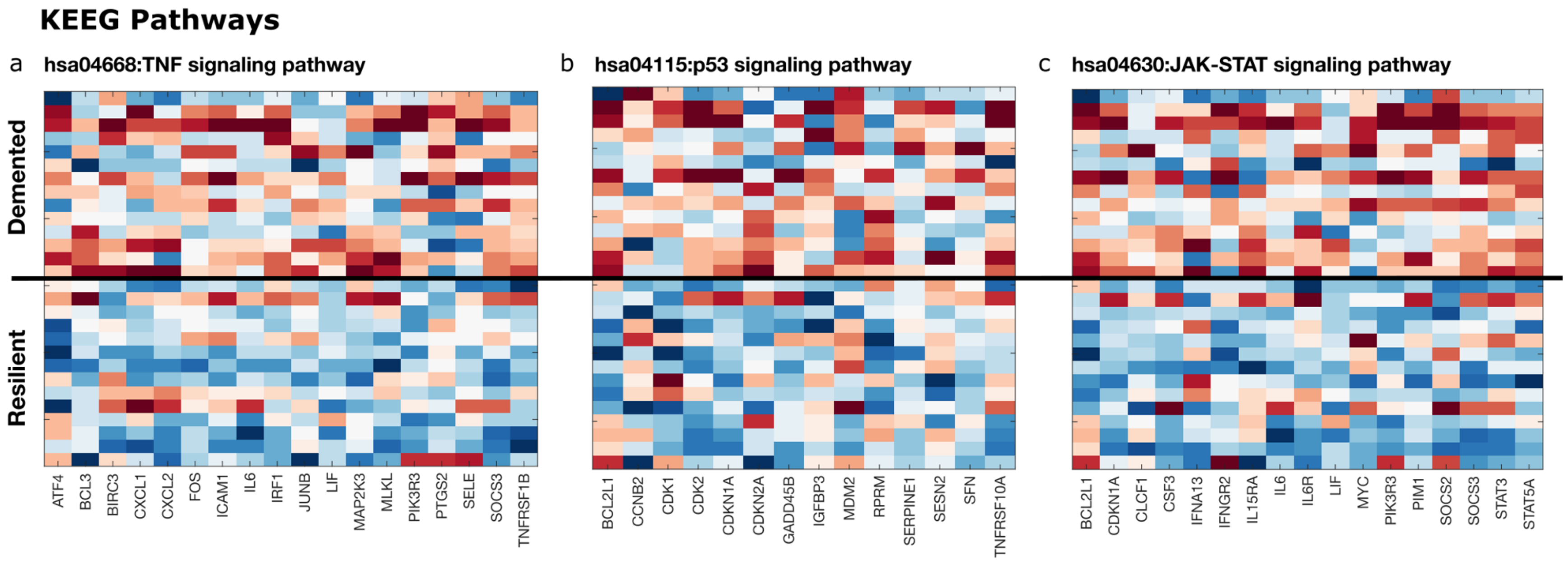


The representative heatmap shows a list of genes that showed significant downregulation in resilient compared to demented AD and were linked to the KEGG pathway, (a) the TNF signaling pathway, (b) the p53 signaling pathway, (c) JAK-STAT signaling pathway.

**eFigure 6.** Workflow for cell type annotation in snRNAseq and identification of microglia cells

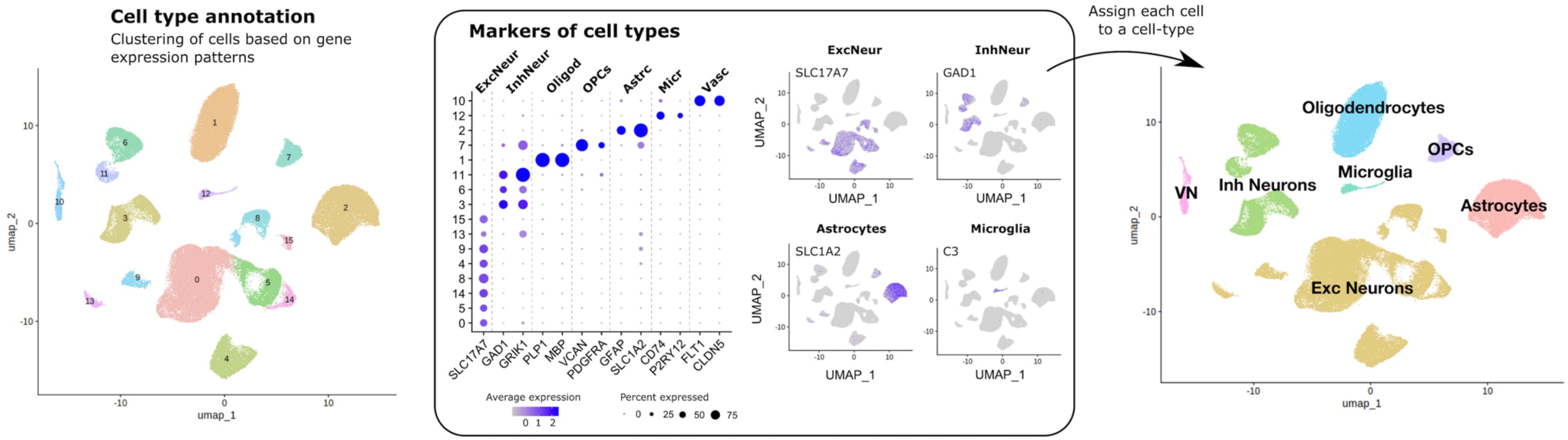


We used the gene expression of each cell to cluster them into different groups. Then we used the expression of known markers/genes to assign each cell to one of 7 mayor cell types in the brain: excitatory neurons, inhibitory neurons, astrocytes, microglia (including monocytes and macrophages), oligodendrocytes, OPCs, and vascular niche (composed of vascular cells, pericytes, SMC, and fibroblasts).

**eFigure 7**. Differential gene expression of IMP-O compared to control brains

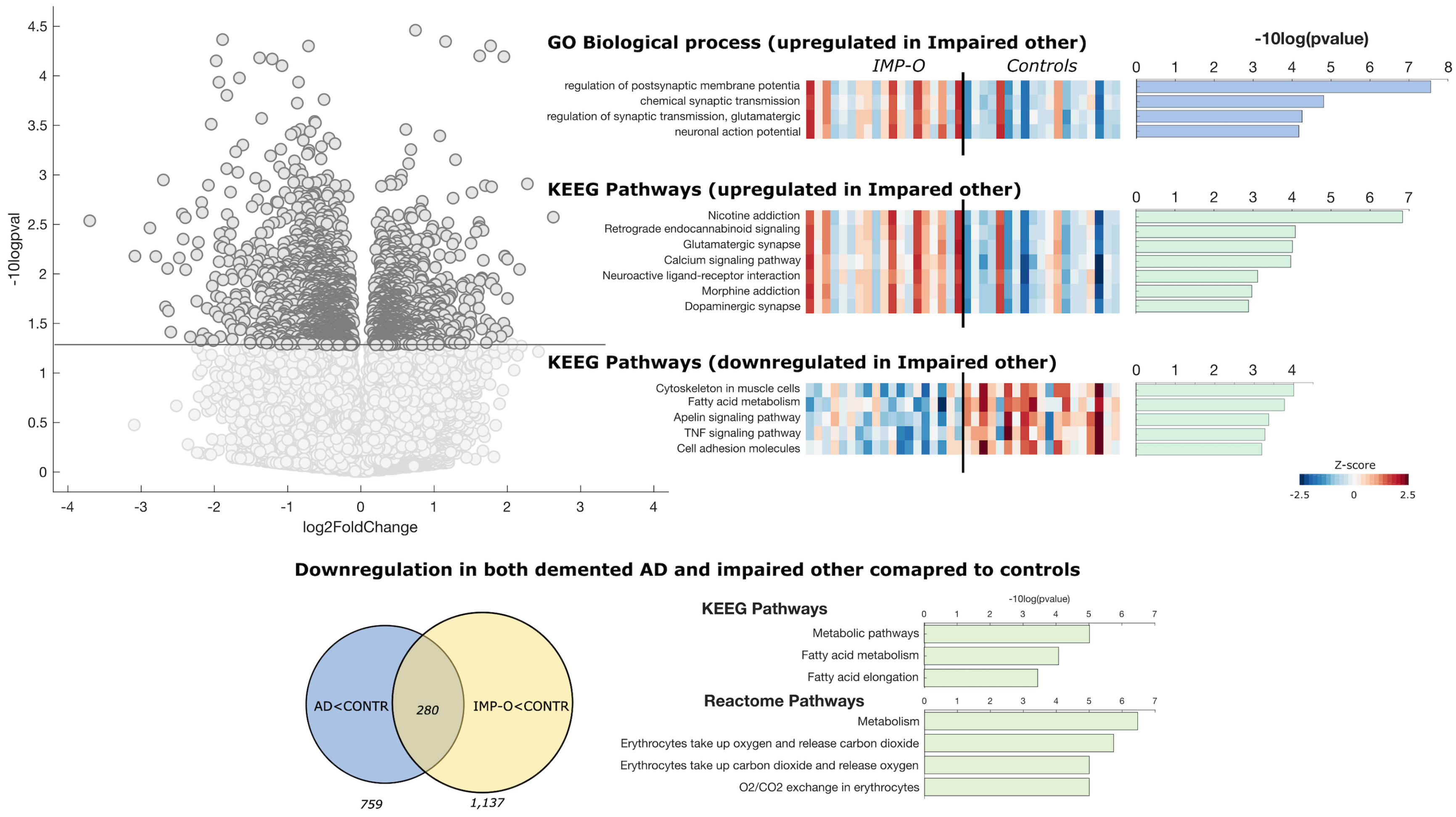


The representative heatmap shows a list of differentially expressed genes in IMP-O compared to control brains and were linkedto (a) upregulated synapse transmission gene ontology process (b) upregulated glutamatergic synapse KEEG pathway and (c) downregulated the TNF signaling KEGG pathways. (d) Venn diagram representing the common downregulated genes in demented AD and IMP-O compared to control cases. The KEGG and reactome pathway suggested that demented AD and IMP-O brains show dysregulated metabolic pathway compared to control brains.
